# Greater reliance on model-free learning in adolescent anorexia nervosa: An examination of dual-system reinforcement learning

**DOI:** 10.1101/2024.01.31.24302097

**Authors:** Carina S. Brown, Sean Devine, A. Ross Otto, Amanda Bischoff-Grethe, Christina E. Wierenga

**Author notes:** **Corresponding Author:** Christina E. Wierenga, Ph.D. Professor of Psychiatry UCSD Eating Disorder Research and Treatment Program UCSD Department of Psychiatry Chancellor Park 4510 Executive Dr., Suite 315 San Diego, CA 92121 http://eatingdisorders.ucsd.edu.

## Abstract

Alterations in learning and decision-making systems are thought to contribute to core features of anorexia nervosa (AN), a psychiatric disorder characterized by persistent dietary restriction and weight loss. Instrumental learning theory identifies a dual-system of habit and goal-directed decision-making, linked to model-free and model-based reinforcement learning algorithms. Difficulty arbitrating between these systems, resulting in an over-reliance on one strategy over the other, has been implicated in compulsivity and extreme goal pursuit, both of which are observed in AN. Characterizing alterations in model-free and model-based systems, and their neural correlates, in AN may clarify mechanisms contributing to symptom heterogeneity (e.g., binge/purge symptoms). This study tested whether adolescents with restricting AN (AN-R; *n* = 36) and binge/purge AN (AN-BP; *n* = 20) differentially utilized model-based and model-free learning systems compared to a healthy control group (HC; *n* = 28) during a Markov two-step decision-making task under conditions of reward and punishment. Associations between model-free and model-based learning and resting-state functional connectivity between neural regions of interest, including orbitofrontal cortex (OFC), nucleus accumbens (NAcc), putamen, and sensory motor cortex (SMC) were examined. AN-R showed higher utilization of model-free learning compared to HC for reward, but attenuated model-free and model-based learning for punishment. In AN-R only, higher model-based learning was associated with stronger OFC-to-left NAcc functional connectivity, regions linked to goal-directed behavior. Greater utilization of model-free learning for reward in AN-R may differentiate this group, particularly during adolescence, and facilitate dietary restriction by prioritizing habitual control in rewarding contexts.

## Introduction

Anorexia nervosa (AN) is a debilitating and potentially life-threatening disorder with one of the highest mortality rates among psychiatric conditions^1–4^. AN typically onsets in adolescence, with at least a third of affected individuals developing a chronic course of illness into adulthood^5^, yet mechanisms contributing to progression of illness are poorly understood. Recent conceptualizations posit a role of habit and goal-directed behavioral systems in the pathogenesis and maintenance of the disorder, although their relative contribution is debated^6–9^. Goal-directed control consists of flexible decision-making, in which the value of specific outcomes can change depending on shifting environmental contingencies. Conversely, habitual control consists of inflexible decision-making, in which choice behavior is difficult to change, even during instances of outcome devaluation^10^.

The central AN symptom of dietary restriction can be described as highly goal-directed, with persistent food restriction in service of long-term rewards (i.e., weight loss) indicating an effortful and goal-driven strategy. Alternatively, the rigidity with which individuals with AN uphold their caloric restriction and the cognitive inflexibility that is observed in their decision-making, might indicate greater use of habit-driven strategies. It has been proposed that what begins as a largely goal-directed pursuit of weight loss, driven by action-outcome learning (e.g., restriction becomes associated with weight loss outcomes), ultimately becomes habitual as repeated actions are overtrained and stimulus-response learning results in specific cues being automatically associated with disorder-relevant actions (e.g., palatable food cues become associated with restriction)^7^. Many individuals with AN also experience episodes of binge eating and purging, behaviors thought to reflect compulsivity (e.g., inflexible, repetitive behavior), and may too reflect greater reliance on habit. Thus, there is a question of how individuals with AN arbitrate between these systems and how this corresponds to restriction versus binge/purge symptomatology.

A compelling computational framework for this dual-system theory of goal-directed and habit decision-making exists in a class of reinforcement learning algorithms known as model-based and model-free learning^11–13^. These algorithms provide instantiations of neural computations that can be behaviorally modeled using variants of multistep Markov reward-based decision-making tasks that have been validated in both human^14^ and animal studies^15^.

Model-based learning algorithms consist of cognitive maps of state-action space that track environmental contingencies, such as transition probabilities when moving from one state to another, and consider these maps when selecting actions^16^. Consequently, this system is behaviorally akin to planning and, although flexible, imposes a considerable computational cost on the decision-maker^17^. The orbitofrontal cortex (OFC), which tracks value computations in response to feedback or shifting motivational and affective states^18–21^, and the ventromedial prefrontal cortex (vmPFC) are implicated in model-based learning^22–25^, with greater intrinsic resting functional connectivity between the medial OFC and nucleus accumbens (Nacc), which tracks prediction errors and anticipated reward and punishment outcomes^26–29^, correlated with greater model-based behavior^24^.

In contrast, model-free learning algorithms are linked to temporal difference learning frameworks, where value estimates based on outcomes are stored across trials and selected actions are dependent on the history of reward receipt, without consideration of environmental contingencies^30^. This system is similar to trial-and-error learning and is not easily adaptable, however it can be quite efficient, akin to habit^31^. Although neural signatures of model-based and model-free learning overlap in the medial prefrontal cortex and the ventral striatum, the putamen has also been implicated in habit behavior during sequential decision-making and outcome devaluation^14,23,32–36^, with putamen-supplementary motor are (SMA) functional connectivity, as well as increased neurite density in the left posterior putamen and right SMA, associated with model-free learning^24^.

Across psychiatric conditions, there is emerging evidence suggesting that disorders of compulsivity (e.g., obsessive-compulsive disorder, substance use) demonstrate deficits in model-based learning during sequential decision-making^25,37–40^. Critically, prior work examining model-based and model-free learning in a mixture of community and clinical samples endorsing symptoms of eating pathology (e.g., compulsive eating), also demonstrate attenuation in model-based learning^25,37–39,41^. Only one such study was conducted in a sample of individuals with full-threshold AN and found evidence of deficits in model-based learning for food and money that were not remediated at weight restoration, reflecting a potential trait behavioral marker of the disorder.

The current study aimed to characterize reliance on model-free and model-based reinforcement learning in AN by clarifying several key issues. First, because AN typically onsets during adolescence and is associated with high risk of chronicity, it is critical to uncover developmental mechanisms that contribute to maintenance of the disorder. Moreover, this developmental period consists of a time frame when model-based learning begins to come online^42^, highlighting a possible treatment target if the relative balance between model-free and model-based systems go awry during initial stages of psychiatric illness. Second, there is heterogeneity in the AN phenotype, with some patients exhibiting only dietary restriction (AN-R) and others also exhibiting binge/purge behaviors (AN-BP). Divergence of symptoms may suggest dissociable deficits in decision-making in individuals with AN-R versus AN-BP. Finally, while most studies have focused exclusively on reward learning, recent research suggests avoidance motivation promotes goal-directed learning^43^, and in OCD, learning bias shifts towards goal-directed control under conditions of loss^44^. Although reward deficits are well-established in AN^45–50^, altered punishment motivation and learning might be particularly salient^51–55^, as AN is characterized by high levels of harm avoidance, exaggerated neural response to losses, and worse treatment outcomes associated with poorer loss-related learning^51,56,57^. However, this may differ by subtype as eating disorders characterized by binge/purge symptoms demonstrate increased limbic-striatal reward response^45–47,50,58,59^, potentially indicating that both subtype and outcome valence may affect utilization of model-based and model-free learning systems.

To elucidate these questions, we conducted a monetary outcomes version of the two-step sequential decision-making task under conditions of reward and punishment in a sample of female adolescents with AN and demographically-matched healthy controls (HC). Participants also completed a separate resting-state functional magnetic resonance imaging scan (rsfMRI) to ascertain whether corticostriatal goal-directed and habit circuits were differentially associated with model-based and model-free learning in AN compared to HC. We hypothesized that adolescents with AN would have attenuated model-based learning compared to HC (especially AN-BP), consistent with increased compulsivity, and greater reliance on model-free learning (especially AN-R), consistent with the notion that dietary restriction is habit-based. With regards to neural functioning, we hypothesized that greater utilization of model-free learning would be linked to stronger functional connectivity between the putamen and SMA, while greater utilization of model-based learning would be linked to stronger functional connectivity between the OFC and NAcc. Finally, we explored whether model-based and model-free learning for reward and punishment were associated with clinical variables.

## METHOD

### Participants

Participants were all female and adolescent, with 36 meeting DSM-5 criteria for restricting type AN (AN-R; age range = 13.67-18.08), 20 meeting DSM-5 criteria for binge-purge type AN (AN-BP; age range = 13.08-18.17), and 28 being demographically matched healthy controls (HC; age range = 13.42-17.83). Approximately 5% of this sample (*n* = 4) completed the reinforcement learning task but did not complete the rsfMRI scan. One subject was excluded due to excessive motion during the rsfMRI scan. Participants included in resting-state analyses included 35 AN-R, 17 AN-BP, and 27 HC. Individuals with AN were recruited from the University of California, San Diego Eating Disorders Treatment and Research Program^60^, and healthy controls were recruited from the community. The study was approved by the University of California San Diego Institutional Review Board, and all participants were provided written informed consent (see Supplement for details regarding participants and assessments).

### Procedure

#### Two-Step Sequential Decision-Making Task

Participants completed a modified two-step sequential decision-making task^14^, which was previously validated in an adolescent sample^42^. They performed the task under conditions of reward, where they either won $1 or won nothing, and punishment, where they either lost $1 or lost nothing (Figure 1A). The order of the conditions was counterbalanced within each group, with each condition consisting of four blocks and a total of 200 trials (see Supplement for task details).

**Figure 1.**
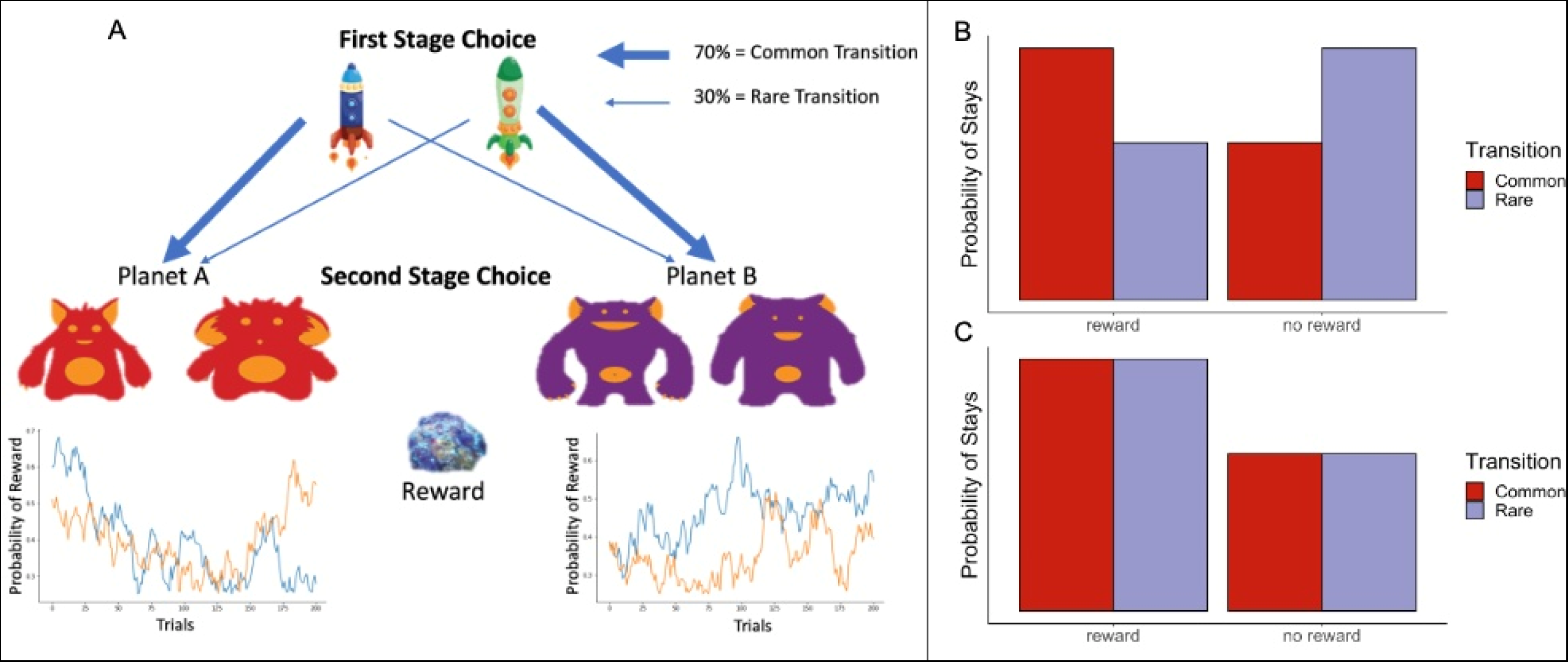
Design of the reward version of the sequential spaceship task based on Decker et al. (2016; A) and sample model-based (B) and model-free (C) behavior. On each trial, participants chose between two spaceships (first-stage choice), which was followed by a probabilistic transition to a red planet or a purple planet. Then participants chose between two aliens (second-stage choice) and were rewarded with space treasure or not. The probability of winning space treasure is presented as a function of trial for each alien. The bar graphs show, for idealized model-free and model-based learners, the probability of making the same choice on the next trial (i.e., a first-stage stay) as a function of the outcome and transition type (common or rare) of the previous trial.

**Figure 2.**
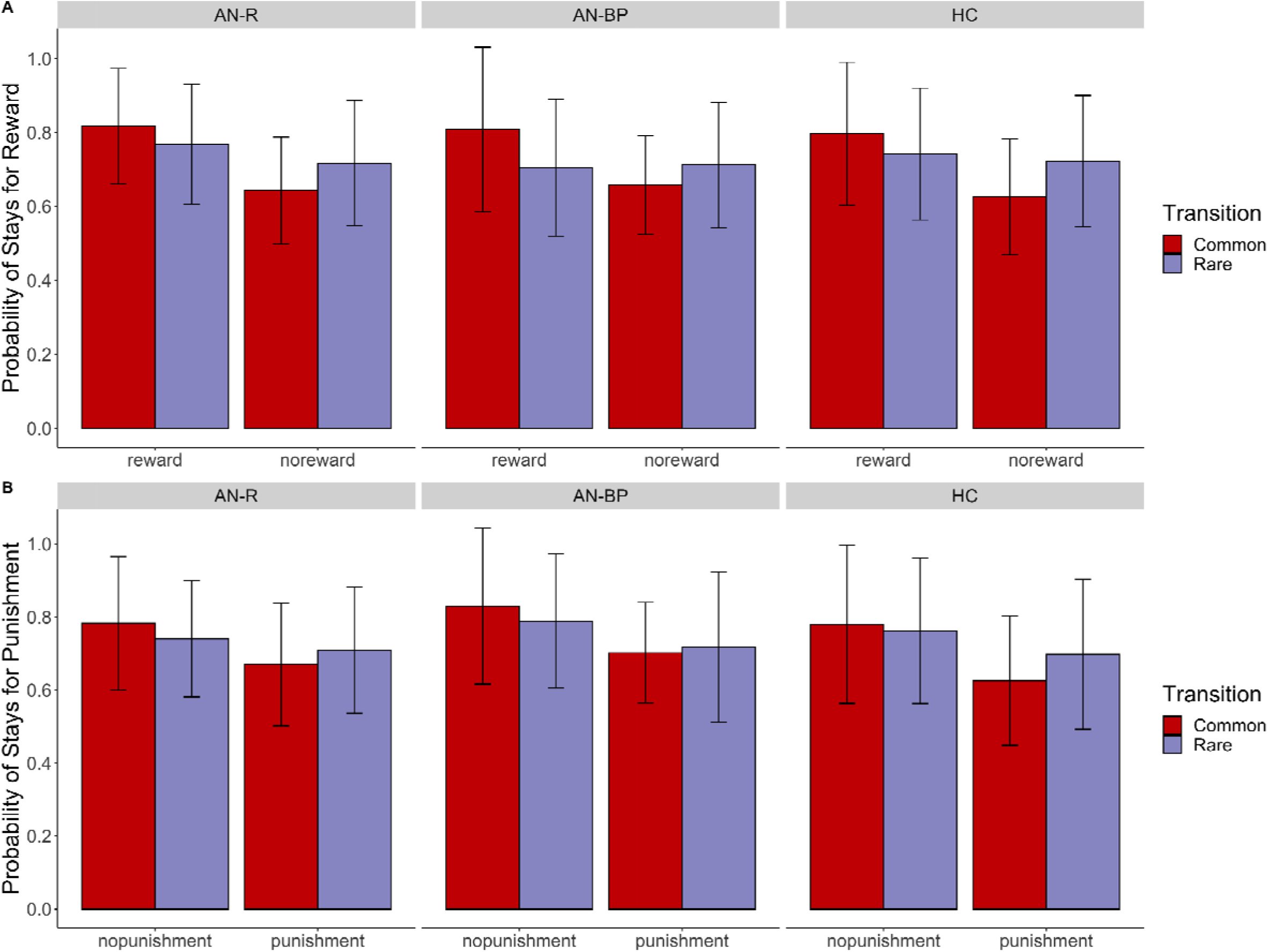
Probability of first-stage stays for the reward (A) and punishment (B) conditions for each group. We visualized the effect of model-based and model-free learning on raw choice data for reward and punishment conditions by plotting stay/switch behavior as a function of outcomes and transition types (i.e., common and rare transitions). The proportion of first-stage stay choices is graphed as a function of outcome of the previous trial for group, separately for trials following common and rare transitions. The error bars represent ±1 SD.

#### Functional Magnetic Resonance Imaging (fMRI)

Resting-state fMRI was acquired separately on a 3.0 T GE MR750 scanner equipped with quantum gradients providing echo planar capability, using a Nova Medical 32 channel head coil (maximum gradient strength: 50 mT/m, slew rate: 200 T/m/s). An eyes-open protocol was implemented, with participants instructed to look at fixation crosshair for the duration. The 1 hour session included: a three-plane localizer; sagittally acquired (0.8 mm slice thickness, FOV=256×240 mm) T1-weighted (MPRAGE PROMO, TE=2.3 ms, flip angle=8o, matrix=320, 2x in-plane acceleration, scan time=7:50) and T2-weighted (3D CUBE, TE=60 ms, variable flip angle, matrix=320, 2x in-plane acceleration, scan time=6:51) whole brain scans for alignment and morphometry; and T2* weighted echo-planar imaging (EPI; 749 volumes, TR= 800 ms, TE=37 ms, flip angle=52°, FOV=180 (RL) x 208 (AP) mm, 72 2 mm thick axial slices, multiband factor=8).

### Statistical Analysis

#### Analysis of Raw Choice Data

Consistent with prior work^14,61^, mixed-effects logistic regression analysis was performed on the raw choice data using *lme4* package (Version 1.1-8)^62^ for the R software environment (Version 4.0.1)^63^. We estimated separate models for reward and punishment conditions. This model predicted first-stage choices (i.e., stay versus switch with respect to the previous response) as a function of previous outcome (i.e., won/lost in previous trial), previous transition type (i.e., rare/common), group, and their interaction. All two- and three-way interactions were defined as fixed effects, with a random intercept and subject-level adjustments to outcome, transition, and their interaction set as random slopes. The lme4 syntax is as follows:

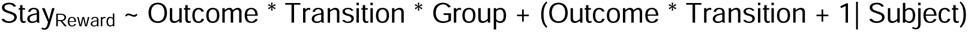

From this analysis, we estimated a model-free effect, approximated by a main effect of previous outcome on stay probability, as well as a model-based effect, approximated by a significant outcome-by-transition interaction.

Percentage of optimal choices was calculated for the second stage decision, specifically the choice between two second-stage stimuli. This was done separately for each condition and for every subject. Means for each group, and for each condition, were subjected to one sample t-tests, comparing to chance (i.e., 50%). Reaction time data for each stage and condition were compared between groups. Results for these analyses are presented in the Supplement (Figures S1-S2).

#### Computational Model

We fit participants’ choice data to a popular reinforcement learning model previously used to model two-stage choice data^14,42,61^. In this “hybrid” model, participants’ choices are theorized to reflect a weighted combination of model-free and model-based learning. This model has been well-validated in a range of psychiatric disorders, with prior work establishing its reliability in a range of psychiatric disorders^37^. The degree to which participants’ responses are governed by principles of model-free or model-based learning is controlled, respectively, by the *β_MF_* and *β_MB_* parameters. Both parameters are bounded at zero and larger values reflect increased model-free or model-basedness.

These model-based and model-free weight parameters were estimated with a hierarchical Bayesian model-fitting technique. To better understand how diagnostic group membership affected model parameters of interest, these parameters were determined on the basis of group membership (akin to regression) estimating coefficients (*γ*) that encoded how these parameters differed between each group (HC, AN-R, AN-BP):

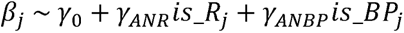

where *is_R* and *is_BP* are dummy-coded variables reflecting whether participant *j* was (or was not) in the AN-R or AN-BP subgroup respectively. Thus, *γ*_0_ encoded model weights in HCs and the other y coefficients tested for differences between HCs and AN subgroups.

To assess the statistical reliability of the observed parameter estimates, a 95% credibility interval (CI) was computed for each parameter. If the CI was wholly situated above or below zero, we determined with 95% confidence that the corresponding parameter was principally positive or negative, respectively. Moreover, we computed Bayesian *p*-values (P), which represent one minus the proportion of the posterior that falls above or below zero (depending on the sign of the median posterior value: below zero if b < 0 and above if b > 0). In line with the traditional interpretation of frequentist *p*-values, Bayesian *p*-values can be interpreted probabilistically as “there is a (P×100) percent chance that the effect is zero or a reversal of the central tendency.” Detailed information about the model can be found in the Supplement.

#### Exploratory analyses with clinical variables

We extracted subject-level random effects from the hierarchical estimation procedure described above to examine the relationship between our computational parameters of interest (*β_MB_* and *β_MF_*) and clinical variables. These random effects estimates were non-linearly transformed before additional analyses, as they were not normally distributed as indicated by Shapiro-Wilk tests (*p*s<.001). To transform the data, we converted the estimates to percentile scores and applied a probit link function, converting to z-scores, which improved normality of the distributions (*p*s>.05). Transformed estimates were used as dependent variables in a series of mixed-effects regression analyses with a group-by-condition-by-measure 3-way interaction term. All analyses controlled for z-scored BMI, IQ, and age as additional covariates. Analyses were conducted and estimated using the R *lme4* package and family-wise Bonferroni-corrected to account for the exploratory nature of the analyses. Mixed-effects linear regression analyses were conducted with the following measures: Eating Disorder Examination (EDE), State-Trait Anxiety Inventory (STAI), Beck Depression Inventory – II (BDI-II), Temperamental Character Inventory – Harm Avoidance (TCI), Sensitivity to Punishment and Reward Questionnaire (SPSRQ), Behavioral Inhibition/Behavioral Activation Scales (BIS/BAS), and Adult Temperament Questionnaire – Effortful Control (ATQ; see Supplement for descriptions). The analysis involving the EDE was only conducted in AN-R and AN-BP, as the majority of HC had total scores of zero.

#### Resting-State Functional Magnetic Resonance Imaging Analysis

Images were processed using scripts developed by the Human Connectome Project freely available online (https://github.com/Washington-University/HCPpipelines<u>). We</u> adapted the HCP quality control metrics^64^, and HCP preprocessing requirements^65^, prior to using the HCP Workbench. Functional images were processed to remove spatial distortion, motion corrected, aligned to the subject’s structural volume, bias field corrected, transformed to standard space, and smoothed using FSL and other tools developed for the HCP protocol. ICA + FIX^66^ was applied to remove spatially specific structured noise, such as motion and transient head movement due to swallowing^67^. Subjects were excluded if any one parameter’s average exceeded 2 standard deviations from the mean to ensure that group differences were not due to motion. One subject was removed from the dataset following preprocessing due to excessive motion.

To investigate whether resting-state functional connectivity is associated with model-based and model-free learning, we conducted an ROI-to-ROI analysis on an *a priori* network. Specifically, we selected regions that have been implicated in both habit and goal-directed learning, including the OFC, NAcc, bilateral putamen, and SMA. Connectivity analyses were conducted in *nilearn*, a python-based brain imaging toolbox (https://nilearn.github.io/index.html). Anatomical ROIs were identified using the Harvard-Oxford Cortical and Subcortical Atlases^68–71^. Timeseries from each ROI were extracted and subjected to ROI-to-ROI analysis. Connectivity values (Fisher z-transformed correlation coefficients) between OFC-NAcc and putamen-SMA dyads were extracted from the connectivity map and entered into mixed-effects linear regression models. All analyses utilized transformed subject-level parameters of interest from the computational model. To examine links between model-based behavior and associated neural regions, model-based estimates (*β_MB_*) were set as the dependent variable with OFC-NAcc connectivity, group, condition, a connectivity-by-group-by-condition 3-way interaction, z-scored age, BMI, and IQ entered as independent variables. We specified the same model for model-free estimates (*β_MF_*) and putamen-SMA connectivity to examine associations between behavioral and neural regions implicated in habit learning. Because these analyses are linked to *a priori* hypotheses about the relationships and direction of relationships, we did not correct for multiple comparisons. Additionally, ROI-to-ROI connectivity was represented using plotting utilities from *nilearn* (Figure 6C).

## RESULTS

### Sample Characteristics

Groups did not differ on age, education, race, ethnicity, IQ, or length of illness (for AN-R and AN-BP only); however, they did differ on BMI, lowest lifetime BMI (for AN-R and AN-BP only), anxiety (STAI-T and STAI-S), depression (BDI), and eating disorder severity (EDE; Table 1).

**Table 1.**
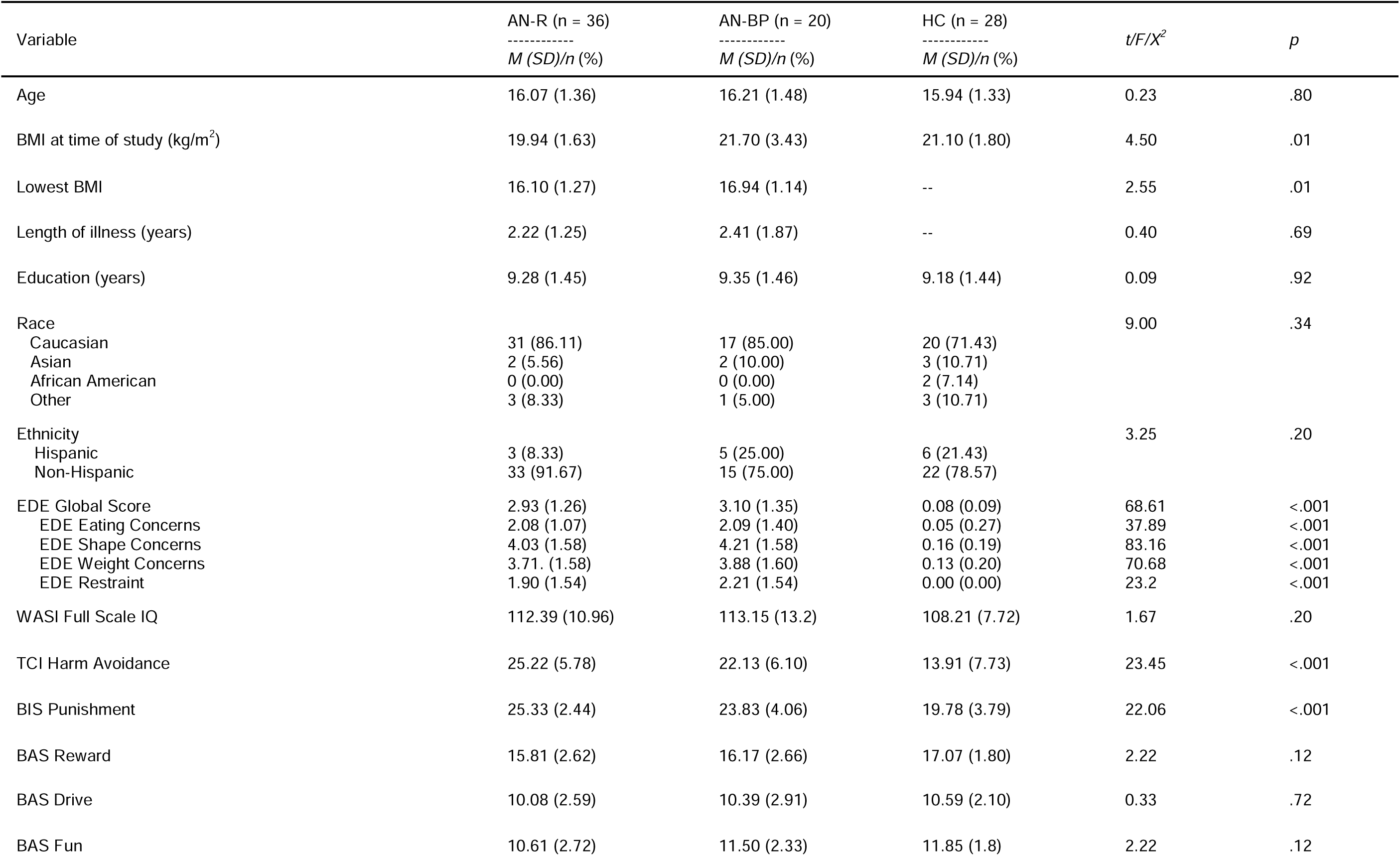
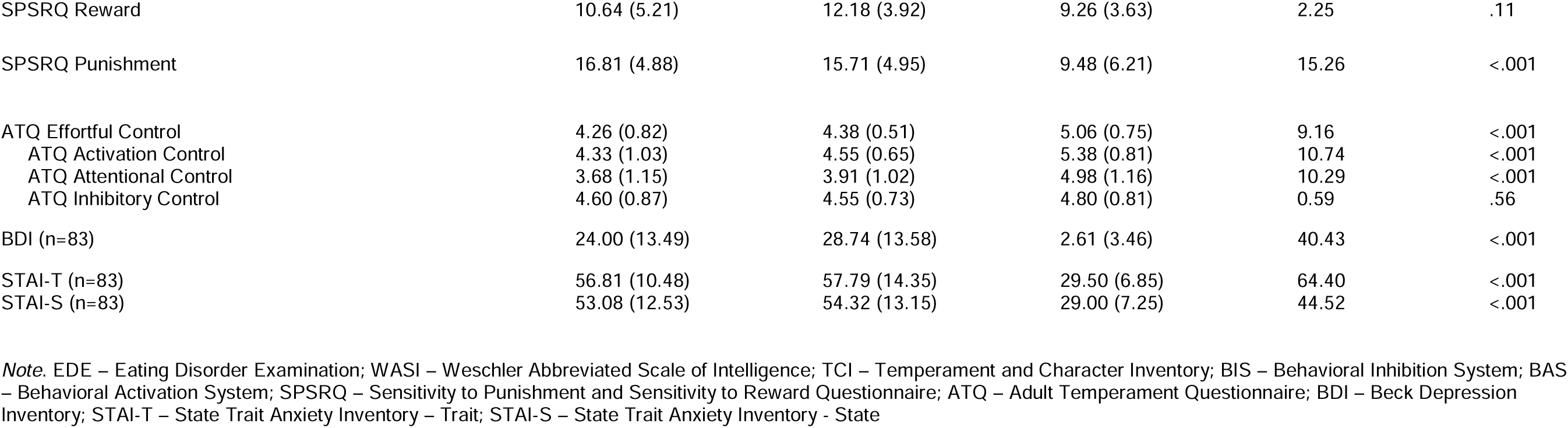
Demographic Characteristics of Sample.

| Variable | AN-R (n = 36)<br>-----<br>*M (SD)/n (%)* | AN-BP (n = 20)<br>-----<br>*M (SD)/n (%)* | HC (n = 28)<br>-----<br>*M (SD)/n (%)* | *t/F/χ<sup>2</sup>* | *p* |
| --- | --- | --- | --- | --- | --- |
| Age | 16.07 (1.36) | 16.21 (1.48) | 15.94 (1.33) | 0.23 | .80 |
| BMI at time of study (kg/m<sup>2</sup>) | 19.94 (1.63) | 21.70 (3.43) | 21.10 (1.80) | 4.50 | .01 |
| Lowest BMI | 16.10 (1.27) | 16.94 (1.14) | -- | 2.55 | .01 |
| Length of illness (years) | 2.22 (1.25) | 2.41 (1.87) | -- | 0.40 | .69 |
| Education (years) | 9.28 (1.45) | 9.35 (1.46) | 9.18 (1.44) | 0.09 | .92 |
| Race |  |  |  | 9.00 | .34 |
| Caucasian | 31 (86.11) | 17 (85.00) | 20 (71.43) |  |  |
| Asian | 2 (5.56) | 2 (10.00) | 3 (10.71) |  |  |
| African American | 0 (0.00) | 0 (0.00) | 2 (7.14) |  |  |
| Other | 3 (8.33) | 1 (5.00) | 3 (10.71) |  |  |
| Ethnicity |  |  |  | 3.25 | .20 |
| Hispanic | 3 (8.33) | 5 (25.00) | 6 (21.43) |  |  |
| Non-Hispanic | 33 (91.67) | 15 (75.00) | 22 (78.57) |  |  |
| EDE Global Score | 2.93 (1.26) | 3.10 (1.35) | 0.08 (0.09) | 68.61 | <.001 |
| EDE Eating Concerns | 2.08 (1.07) | 2.09 (1.40) | 0.05 (0.27) | 37.89 | <.001 |
| EDE Shape Concerns | 4.03 (1.58) | 4.21 (1.58) | 0.16 (0.19) | 83.16 | <.001 |
| EDE Weight Concerns | 3.71 (1.58) | 3.88 (1.60) | 0.13 (0.20) | 70.68 | <.001 |
| EDE Restraint | 1.90 (1.54) | 2.21 (1.54) | 0.00 (0.00) | 23.2 | <.001 |
| WASI Full Scale IQ | 112.39 (10.96) | 113.15 (13.2) | 108.21 (7.72) | 1.67 | .20 |
| TCI Harm Avoidance | 25.22 (5.78) | 22.13 (6.10) | 13.91 (7.73) | 23.45 | <.001 |
| BIS Punishment | 25.33 (2.44) | 23.83 (4.06) | 19.78 (3.79) | 22.06 | <.001 |
| BAS Reward | 15.81 (2.62) | 16.17 (2.66) | 17.07 (1.80) | 2.22 | .12 |
| BAS Drive | 10.08 (2.59) | 10.39 (2.91) | 10.59 (2.10) | 0.33 | .72 |
| BAS Fun | 10.61 (2.72) | 11.50 (2.33) | 11.85 (1.8) | 2.22 | .12 |
| SPSRQ Reward | 10.64 (5.21) | 12.18 (3.92) | 9.26 (3.63) | 2.25 | .11 |
| SPSRQ Punishment | 16.81 (4.88) | 15.71 (4.95) | 9.48 (6.21) | 15.26 | <.001 |
| ATQ Effortful Control | 4.26 (0.82) | 4.38 (0.51) | 5.06 (0.75) | 9.16 | <.001 |
| ATQ Activation Control | 4.33 (1.03) | 4.55 (0.65) | 5.38 (0.81) | 10.74 | <.001 |
| ATQ Attentional Control | 3.68 (1.15) | 3.91 (1.02) | 4.98 (1.16) | 10.29 | <.001 |
| ATQ Inhibitory Control | 4.60 (0.87) | 4.55 (0.73) | 4.80 (0.81) | 0.59 | .56 |
| BDI (n=83) | 24.00 (13.49) | 28.74 (13.58) | 2.61 (3.46) | 40.43 | <.001 |
| STAI-T (n=83) | 56.81 (10.48) | 57.79 (14.35) | 29.50 (6.85) | 64.40 | <.001 |
| STAI-S (n=83) | 53.08 (12.53) | 54.32 (13.15) | 29.00 (7.25) | 44.52 | <.001 |

*Note.* EDE – Eating Disorder Examination; WASI – Weschler Abbreviated Scale of Intelligence; TCI – Temperament and Character Inventory; BIS – Behavioral Inhibition System; BAS – Behavioral Activation System; SPSRQ – Sensitivity to Punishment and Sensitivity to Reward Questionnaire; ATQ – Adult Temperament Questionnaire; BDI – Beck Depression Inventory; STAI-T – State Trait Anxiety Inventory – Trait; STAI-S – State Trait Anxiety Inventory - State

### Raw Choice Data

Model-based and model-free learning were approximated using raw choice data by examining probabilities of first-stage stay behavior. Specifically, increased probabilities of stay versus switch following second-stage wins served as a measure of model-free behavior. Increased probabilities of stay versus switch following wins during common transitions, and losses during rare transitions, served as a measure of model-based behavior. To visualize differences in these systems^41^, we computed difference scores based on prior work by Eppinger et al. (2017). The model-based difference score was taken as: (common/win + rare/lose) – (common/lose + rare/win). The model-free difference score was taken as: (common/win+ rare/win) – (common/lose + rare/lose). These values are plotted as a function of group and condition (Figure 3).

**Figure 3.**
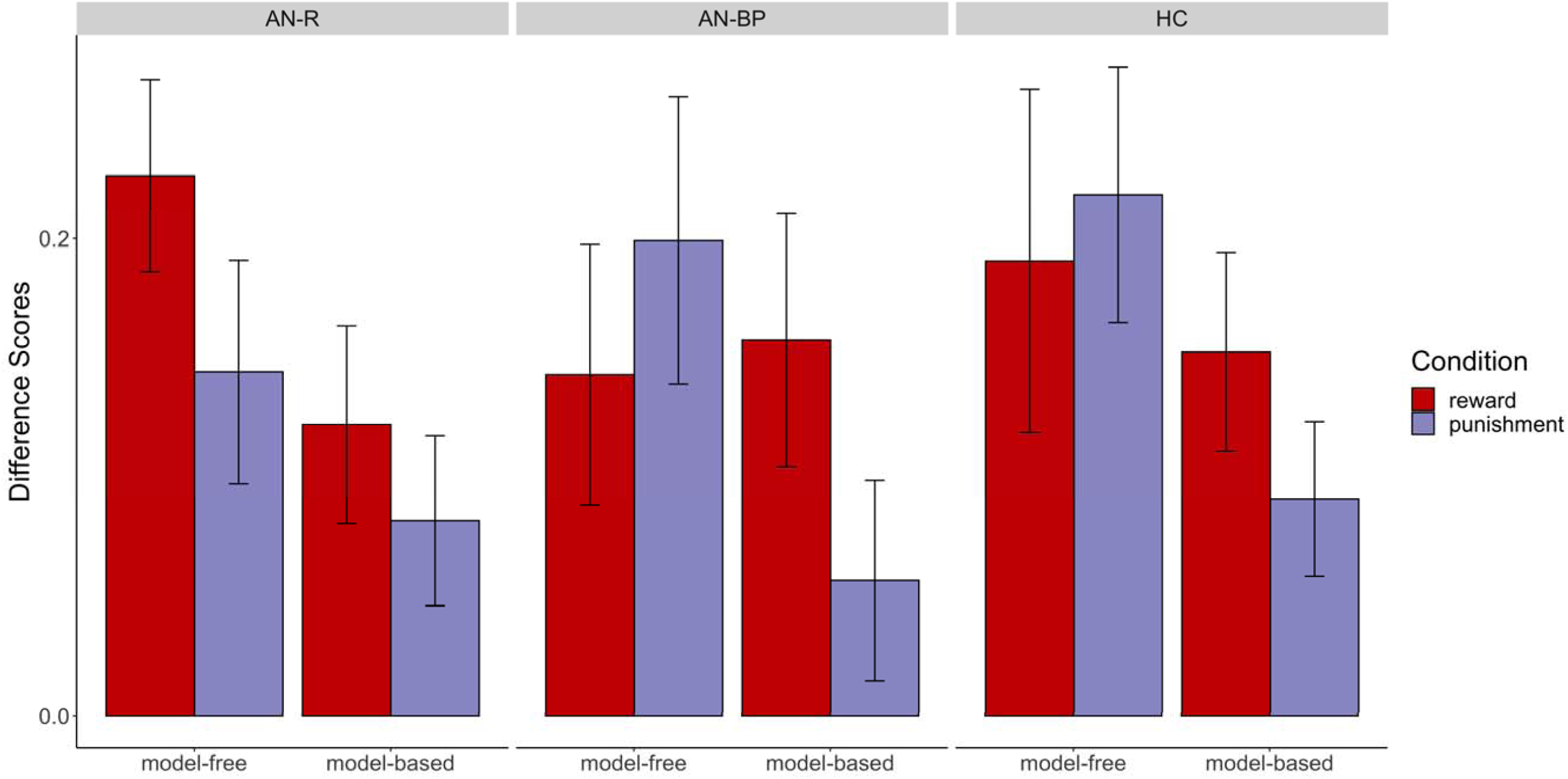
Index of the model-based and model-free effects by condition and group based on first-stage stay probabilities. The model-based difference score was taken as: (common/win + rare/lose) – (common/lose + rare/win). The model-free difference score was taken as: (common/win+ rare/win) – (common/lose + rare/lose). Error bars represent ±1 SEM.

Results from the mixed-effects logistic regression analysis indicate that for reward, there was a significant main effect of outcome (B = 1.27, SE = 0.27, *p* = <.001) and an outcome-by-transition interaction (B = −1.16, SE = 0.32, *p* = <.001), but no significant group differences (Table 2). Punishment was similar, with a main effect of outcome (B = 1.15, SE = 0.23, *p* = <.001) and an outcome-by-transition interaction (B = −0.69, SE = 0.24, *p* = .003), but again no significant group differences (Table 2). An omnibus mixed-effects logistic regression analysis also including condition revealed similar results, with no group differences (see Supplement). Results therefore support mixed usage of model-free and model-based learning across groups, with computational model results below offering a more granular understanding of group differences in the relative balance of these systems.

**Table 2.**
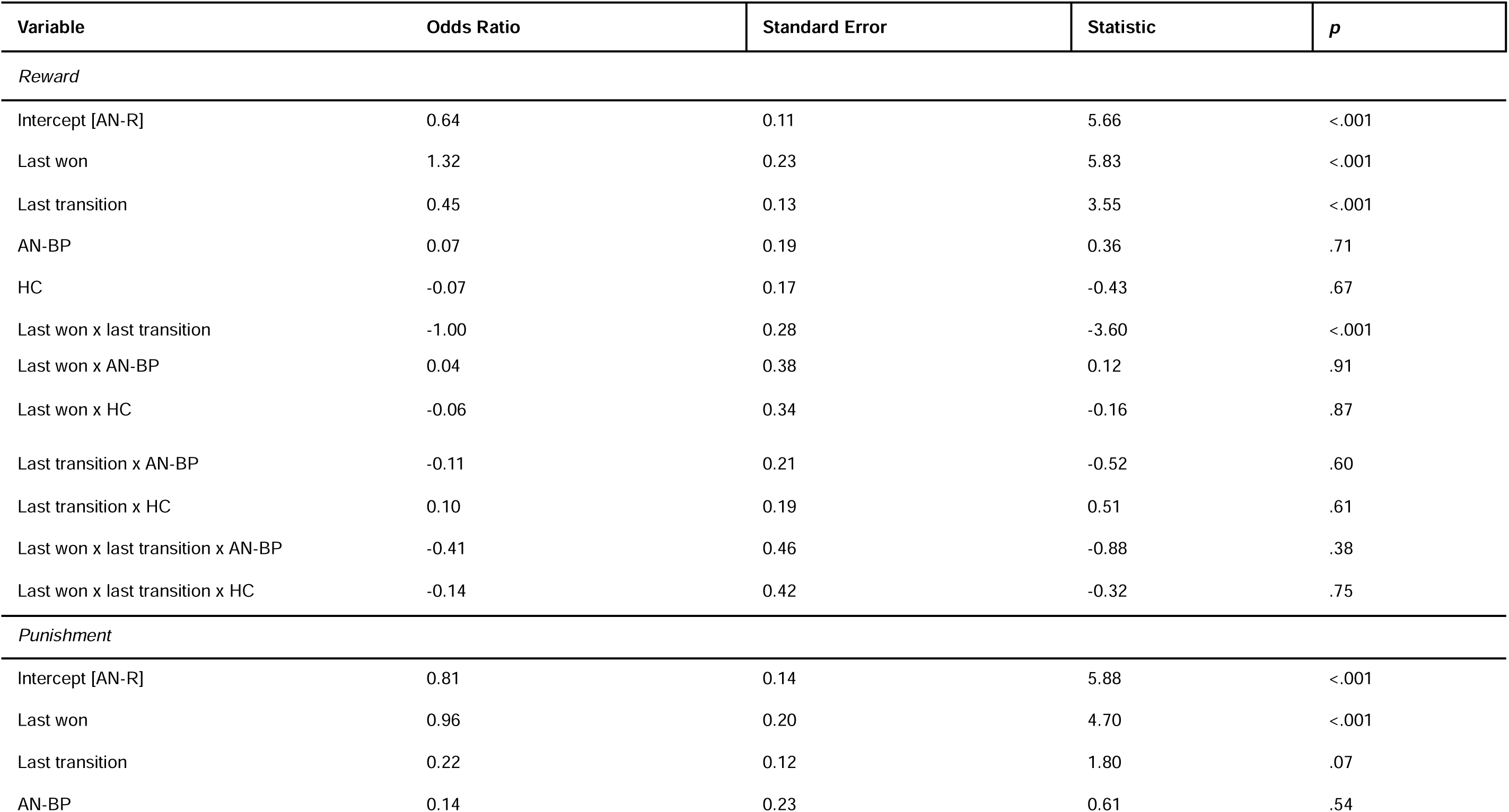
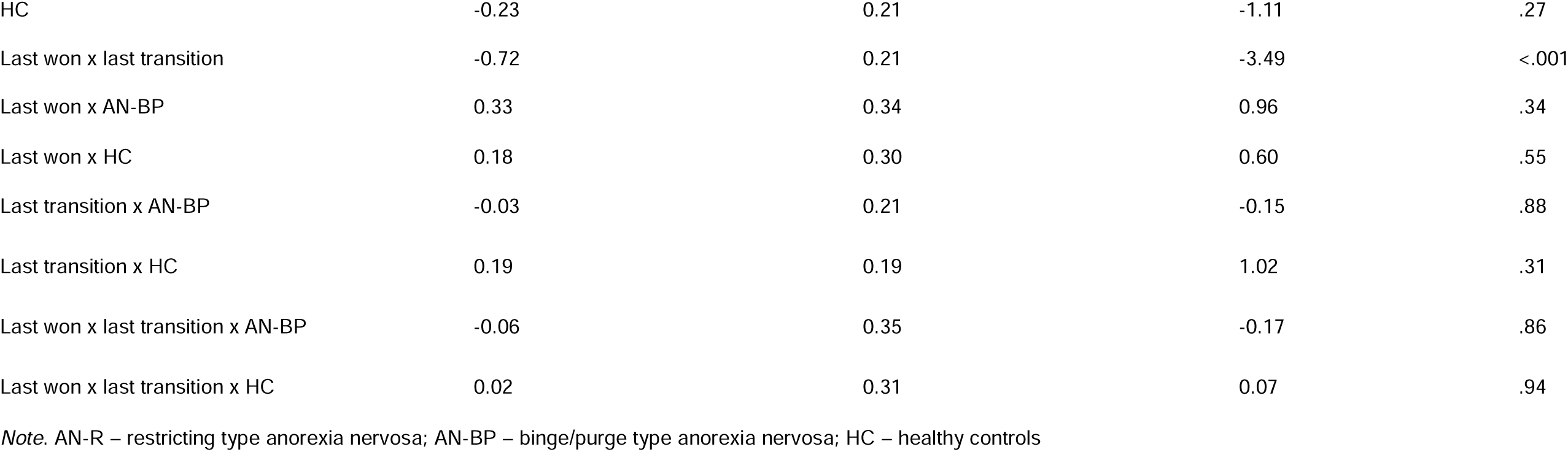
Raw Choice Data Mixed Effects Logistic Regression (By Condition)

| Variable | Odds Ratio | Standard Error | Statistic | *p* |
| --- | --- | --- | --- | --- |
| *Reward* |  |  |  |  |
| Intercept [AN-R] | 0.64 | 0.11 | 5.66 | <.001 |
| Last won | 1.32 | 0.23 | 5.83 | <.001 |
| Last transition | 0.45 | 0.13 | 3.55 | <.001 |
| AN-BP | 0.07 | 0.19 | 0.36 | .71 |
| HC | -0.07 | 0.17 | -0.43 | .67 |
| Last won x last transition | -1.00 | 0.28 | -3.60 | <.001 |
| Last won x AN-BP | 0.04 | 0.38 | 0.12 | .91 |
| Last won x HC | -0.06 | 0.34 | -0.16 | .87 |
| Last transition x AN-BP | -0.11 | 0.21 | -0.52 | .60 |
| Last transition x HC | 0.10 | 0.19 | 0.51 | .61 |
| Last won x last transition x AN-BP | -0.41 | 0.46 | -0.88 | .38 |
| Last won x last transition x HC | -0.14 | 0.42 | -0.32 | .75 |
| *Punishment* |  |  |  |  |
| Intercept [AN-R] | 0.81 | 0.14 | 5.88 | <.001 |
| Last won | 0.96 | 0.20 | 4.70 | <.001 |
| Last transition | 0.22 | 0.12 | 1.80 | .07 |
| AN-BP | 0.14 | 0.23 | 0.61 | .54 |
| HC | -0.23 | 0.21 | -1.11 | .27 |
| Last won x last transition | -0.72 | 0.21 | -3.49 | <.001 |
| Last won x AN-BP | 0.33 | 0.34 | 0.96 | .34 |
| Last won x HC | 0.18 | 0.30 | 0.60 | .55 |
| Last transition x AN-BP | -0.03 | 0.21 | -0.15 | .88 |
| Last transition x HC | 0.19 | 0.19 | 1.02 | .31 |
| Last won x last transition x AN-BP | -0.06 | 0.35 | -0.17 | .86 |
| Last won x last transition x HC | 0.02 | 0.31 | 0.07 | .94 |

*Note.* AN-R – restricting type anorexia nervosa; AN-BP – binge/purge type anorexia nervosa; HC – healthy controls

### Computational Model

The reinforcement learning model consisted of a combination of model-based and model-free weights (*β_MB_* and *β_MF_*, respectively) that control the relative contribution of model-free and model-based updating on choice behavior. Differences between HC and AN subgroups were encoded in regression coefficients (*γ*_0_, *γ_ANR_*, and *γ_ANBP_*) for each freely estimated parameter, with HC being set as the reference group (*γ*_0_): if these coefficients differed from zero, then diagnostic subgroups differed on these parameters from HC. To compare AN subgroups to each other, we reconstructed subgroup posteriors and directly compared these posterior distributions to each other: AN-R = *γ*_0_ + *γ_ANR_* and AN-BP = *γ*_0_ + *γ_ANBP_*. Posterior distributions of transformed *γ* weights are plotted in Figures 4-5. Below, we report average y values, 95% credibility intervals, and Bayesian P-values (see Method).

**Figure 4.**
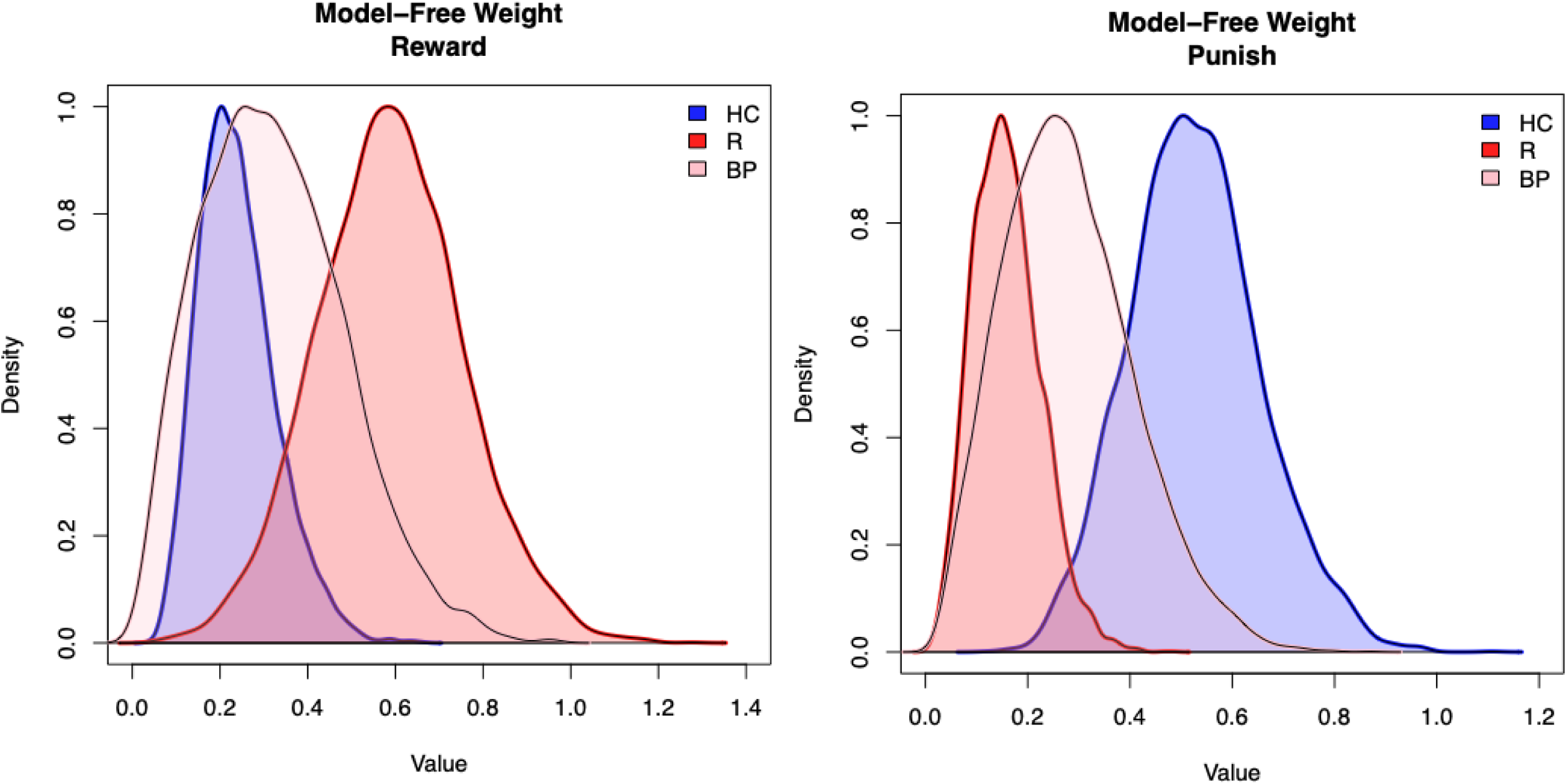
Group-level posterior distributions of model-free weights for reward (left) and punishment (right). Values have been log transformed to display estimates on the correct scale. R – restricting subtype, BP – binge/purge subtype, HC – healthy control

**Figure 5.**
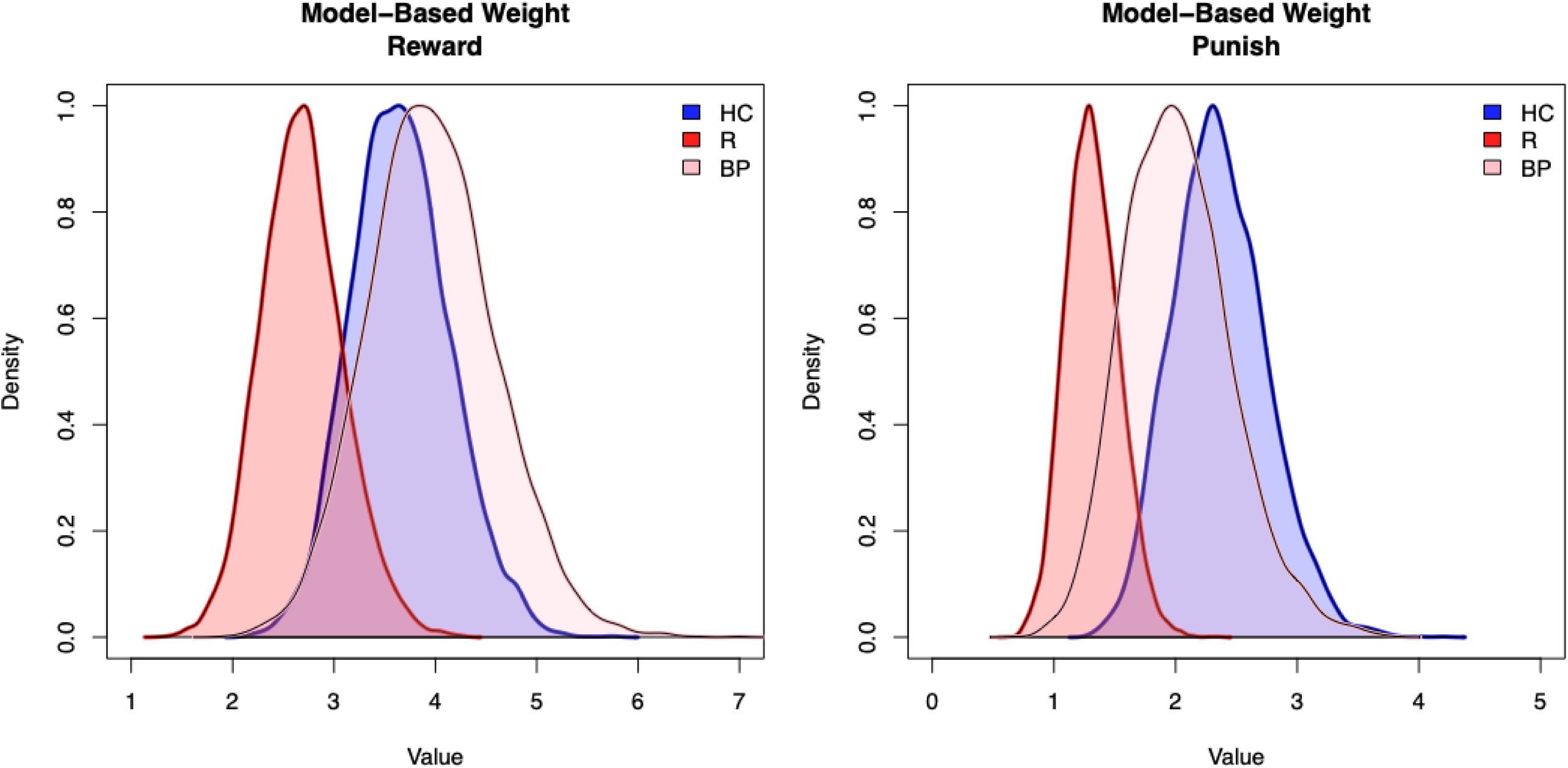
Group-level posterior distributions of model-based weights for reward (left) and punishment (right). Values have been log transformed to display estimates on the correct scale. R – restricting subtype, BP – binge/purge subtype, HC – healthy control

**Figure 6.**
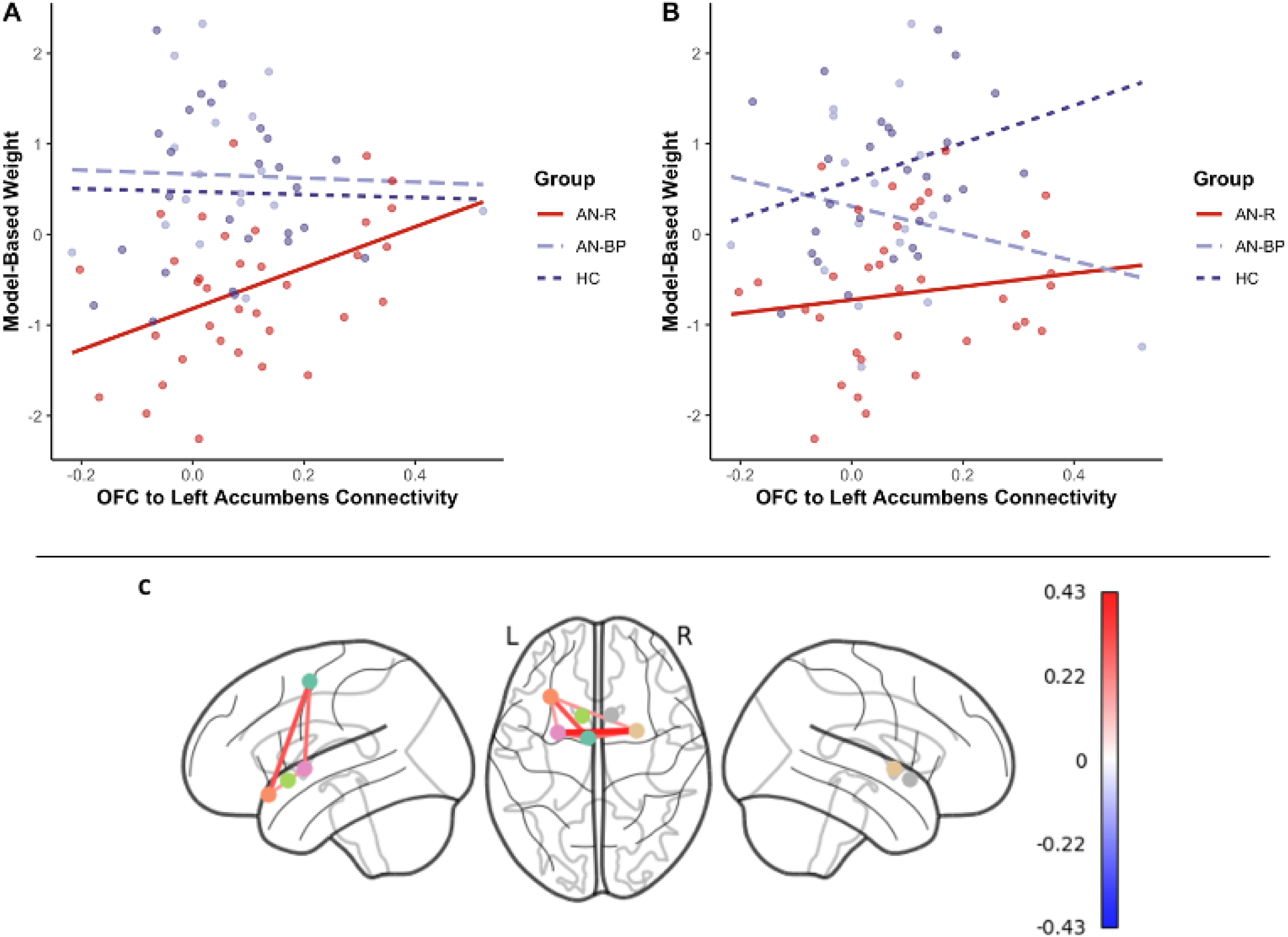
Resting-state functional connectivity results, including associations between model-based weights and OFC-to-left NA connectivity for reward (A) and punishment (B). Covariance matrix for ROI-to-ROI connectivity are visualized as a functional connectome, with edges representing the strength of the correlation between nodes (C). Reinforcement Learning in Anorexia Nervosa

In the reward condition, AN-R had significantly higher model-free learning compared to HC (y_ANR_ = 0.92, *P* = .03, 95%CI[−0.02, 1.82]) and lower model-based learning at a trend level (y_ANR_ = −0.31, *P =* .05, 95%CI[−0.70, 0.07]). In the punishment condition, AN-R displayed significantly attenuated model-free learning (y_ANR_ = −1.26, *P* = .002, 95%CI[−2.38, −0.38]), and model-based learning compared to HC (y_ANR_ = −0.58, *p* = .004, 95%CI[−1.01, −0.16]). AN-BP also had attenuated model-free learning for punishment compared to HC at a trend level (y_ANBP_ = −0.72, *p* = .07, 95%CI[−1.99, 0.21]). Overall, AN-R showed greater reliance on model-free learning for the reward condition, but deficits in both model-free and model-based learning for the punishment condition. Conversely, AN-BP only displayed trend-level deficits in model-free learning for punishment relative to HC. These results are displayed in Tables 3 and 4. Estimated y weights for the remaining freely estimated model parameters (*α*, *β*_2_, *p*) are included in Table S2-S4.

**Table 3.** Latent Regression Coefficients on Model-Free Weights.

| Parameter | Mean Posterior | *p* | CI[2.5%] | CI[97.5%] |
| --- | --- | --- | --- | --- |
| *Reward* |  |  |  |  |
| $\gamma_0$ [HC] | -1.49 | 0 | -2.30 | -0.83 |
| $\gamma_1$ [AN-R vs HC] | 0.92 | .03 | -0.02 | 1.82 |
| $\gamma_2$ [AN-BP vs HC] | 0.20 | .34 | -1.24 | 1.30 |
| *Punishment* |  |  |  |  |
| $\gamma_0$ [HC] | -0.67 | <.001 | -1.23 | -0.23 |
| $\gamma_1$ [AN-R vs HC] | -1.26 | .002 | -2.38 | -0.38 |
| $\gamma_2$ [AN-BP vs HC] | -0.72 | .07 | -1.99 | 0.22 |

*Note.* AN-R – restricting type anorexia nervosa; AN-BP – binge/purge type anorexia nervosa; HC – healthy controls.

**Table 4.** Latent Regression Coefficients on Model-Based Weights.

| Parameter | Mean Posterior | *p* | CI[2.5%] | CI[97.5%] |
| --- | --- | --- | --- | --- |
| *Reward* |  |  |  |  |
| $\gamma_0$ [HC] | 1.29 | 0 | 1.02 | 1.54 |
| $\gamma_1$ [AN-R vs HC] | -0.31 | .05 | -0.70 | 0.07 |
| $\gamma_2$ [AN-BP vs HC] | 0.08 | .34 | -0.32 | 0.47 |
| *Punishment* |  |  |  |  |
| $\gamma_0$ [HC] | 0.85 | <.001 | 0.52 | 1.15 |
| $\gamma_1$ [AN-R vs HC] | -0.58 | .004 | -1.01 | -0.16 |
| $\gamma_2$ [AN-BP vs HC] | -0.17 | .25 | -0.68 | 0.34 |

*Note.* AN-R – restricting type anorexia nervosa; AN-BP – binge/purge type anorexia nervosa; HC – healthy controls. Mean posterior estimates are not transformed.

### Associations with Resting-State Functional Connectivity

We extracted rsfMRI functional connectivity coefficients from subject-level connectivity maps for OFC-NAcc and SMA-putamen pairs. There were no group differences in connectivity between these ROIs, controlling for z-scored age, BMI, and IQ. There were no significant effects between SMA-putamen connectivity and *β_MF_*, including no main effect of connectivity, no connectivity-by-group interaction, and no connectivity-by-condition interaction. For OFC-left NAcc connectivity and *β_MB_*, there was a significant main effect of connectivity for AN-R (B = 2.15, SE = 0.92, *p* = .02), but no significant main effects for any other group. There were also no significant connectivity-by-group interactions, nor were there connectivity-by-condition interactions. Because reward was the reference level for the condition variable, a main effect of connectivity indicated that higher reward *β_MB_* was associated with stronger resting-state connectivity between the OFC-left NAcc in AN-R only (Figure 6A-6B). To explore this relationship within the AN-R group, we conducted a post-hoc linear regression analysis predicting reward *β_MB_* and specifying an OFC-left NAcc connectivity and EDE Global Score interaction, controlling for the same covariates. The main effect of connectivity (B = 7.53, SE = 2.07, *p* = .001) and the interaction term (B = −1.92, SE = 0.70, *p* = .01) were significant, showing that the more positive the relationship between reward *β_MB_* and connectivity, the lower the EDE Global Score (Figure S7).

### Associations with Clinical Variables

Consistent with prior work^41^, there were no significant associations between *β_MB_* or *β_MF_* and symptom measures (BMI, EDE, BDI, or STAI) after family-wise correction. We also did not find significant associations with TCI Harm Avoidance, SPSRQ Reward, SPSRQ Punishment, BIS, or BAS Reward Responsivity. However, when examining *β_MB_* and ATQ Effortful Control, there was a significant Effortful Control-by-condition-by-group interaction, suggesting differences in the directionality of both the effect between AN-BP and AN-R and the effect between conditions for AN-BP, detailed in the Supplement (Table S8, Figure S8).

## Discussion

This study investigated, amongst subtypes of adolescent AN and HC, alterations in model-based and model-free learning under conditions of reward and punishment, as well as associated links to corticostriatal functional connectivity. Overall, AN-R showed greater reliance on model-free learning for reward, but decreased utilization of both model-free and model-based learning for punishment. Moreover, in AN-R only, higher model-based learning for reward was associated with stronger functional connectivity between regions implicated in goal-directedness, and this was associated with lower eating disorder severity. In contrast, adolescents with AN-BP displayed trend-level deficits in model-free learning for punishment, as well as a negative relationship between effortful control and model-based learning for reward, but no other significant behavioral or neural differences when compared to HC or AN-R.

Together, results suggest substantive differences in the balance between model-free and model-based learning between AN-R and HC across conditions, as well as differentiation between AN subtypes primarily under reward. Such alterations in adolescent AN-R may reflect over-reliance on inflexible, habit-based reward learning and inadequate utilization of flexible, goal-directed learning.

Parameter estimates from the reinforcement learning model indicate that AN-R displayed patterns of model-free and model-based learning similar to those observed in OCD^40^. There is accumulating evidence that disorders characterized by compulsivity (e.g., OCD, substance use) rely less on model-based learning during sequential reward-based decision-making tasks^25,37–39,41,44^, indicating that this learning bias may be a transdiagnostic feature of psychiatric conditions characterized by cognitive or behavioral perseveration. Of note, greater utilization of model-free learning in our AN-R sample may be aligned with etiological models purporting that dietary restriction is initially maintained by operant conditioning (e.g., reinforced by reward of weight loss and praise) that becomes a classically conditioned habit over time^8,73,74^ and raises the question of whether a tendency towards reward-based model-free learning may reflect a premorbid susceptibility factor that predisposes the early development of dietary habits. This hypothesis is aided by the positive relationship between model-based learning and OFC-to-left NAcc functional connectivity in AN-R only, potentially indicating that heightened neural synchrony in reward circuitry is connected to greater reward-based goal pursuit. Prior studies have reported disturbances in reward-related functional architecture in AN^75,76^, particularly in ventral and dorsal frontostriatal networks, that support etiological theories implicating alterations in reward functioning and over-reliance on habit^7,73,77,78^. Our post-hoc analysis further clarified that a stronger, positive association between OFC-to-left NAcc functional connectivity and model-based learning was linked to less severe eating pathology, providing potential evidence of a brain-based protective marker, though this was exploratory. If true, this would suggest that learning systems contributing to decision-making under reward and corresponding neural circuitry may be of particular clinical importance^79^.

While behavioral patterns present for AN-R are relatively consistent with prior work in psychiatric samples^25,37–39,41,44^, the lack of difference between AN-BP and HC was unexpected. A prior study showed that AN displayed attenuated model-based learning compared to HC, with this effect being largely driven by AN-BP^41^. Developmental factors and/or illness stage may have contributed to our findings; indeed, prior research demonstrates that utilization of model-based learning can change over the course of development, with model-based systems coming online during adolescence and strengthening into adulthood^42,80–83^. Further, higher compulsivity at baseline in healthy adolescents is associated with diminished strengthening of model-based control at a later time point, suggesting that maturation of this system can be hijacked by compulsive psychological features^82^. It is therefore possible that adolescent AN-R reflects delayed maturation of these systems and AN-BP may not show signs of deficiency at this stage, but may fail to improve as a function of age. Longitudinal studies are needed to resolve this issue.

Our results also suggest that AN-R and AN-BP are behaviorally dissociable, particularly in their responses to reward learning. AN-R utilized greater model-free learning for reward, while AN-BP did not significantly differ from HC in either learning system. Exploratory analyses with clinical variables supported the distinction between subtypes as well, with greater reward-related model-based learning in AN-BP linked to lower levels of adaptive effortful control (i.e., self-regulation), while AN-R displayed a weaker, positive relationship. One possible explanation for this distinction may be differences in saliency of reward. It is postulated that AN-BP is characterized by greater sensitivity to reward compared to AN-R, with some studies corroborating this hypothesis^53,84,85^, and others contradicting it^52,54^. In the current study, self-reported sensitivity to reward is intact in the AN sample and is not differentiated by subtype, but utilization of model-free/model-based learning to make decisions in a rewarding context is.

Model-based control has high computational expense, making it an effortful choice. If individuals with AN-BP are indeed more sensitive to reward, it is possible that positive outcomes (e.g., monetary gain, palatable food consumption) have higher motivational and incentive salience, leading to use of a cognitively demanding strategy even in the context of lower trait self-regulatory control. This is consistent with findings that adults with bulimic-type symptoms are more willing to work for reward as demonstrated by a higher breakpoint during a progressive ratio task^86^, and prior research demonstrating that model-based learning can be modulated by stress and cognitive control^61,87^.

It is worth noting that the results we observed in the punishment condition were not consistent with our hypotheses. Despite greater self-reported sensitivity to punishment (i.e., subjective responsivity and/or avoidance of aversive consequences), AN-R displayed reductions in both model-based and model-free learning compared to HC, raising a question of how this group ultimately made choices if they were relatively deficient in both systems during punishment. Notably, this is in contrast to prior work suggesting increased model-based learning for punishment, which has been observed in OCD^40^, serotonin-depletion^88^, and in healthy adolescents and adults^80^. We previously reported lower learning rates for both reward and punishment in a predominantly young adult sample of women with AN-R; however, worse learning on punishment trials was associated with poorer outcome, suggesting that these individuals may have particular difficulty modifying expectations when punishment is possible.

Whether this relates to active avoidance of aversive outcomes or cognitive inflexibility is difficult to determine, although in the current study, the punishment inverse temperature parameter (*β*_2_) was significantly lower in AN-R compared to HC, which indicates more exploration of choices and less exploitation of choice valuation during the second stage (Table S3). Alternatively, AN-R may have utilized another learning system or strategy (e.g., successor representation) for the punishment condition, although more research is needed to explore this hypothesis.

This study was the first to investigate model-based/model-free learning in adolescent samples of AN-R, AN-BP, and HC. Strengths of this work include a well-characterized adolescent AN sample, use of a well-validated two-step decision-making task with monetary outcomes to avoid symptom provocation, modulation of outcome valence, use of a well-validated computational model to elicit latent learning features, and the inclusion of rsfMRI to examine relationships between cognitive and neural features of learning. Results add to growing literature on the arbitration between these learning systems in psychopathology and provide greater insight into not only earlier stages of development and disease progression, but also valence-specific effects that may be of particular importance in development and maintenance of AN.

### Limitations

Despite these strengths, limitations of this study highlight directions for future work. Our sample included fewer participants with AN-BP compared to AN-R and HC, and both weight-restored and underweight adolescents with AN were included. Additionally, neural data only consisted of rsfMRI scans, with a limited ability to examine task-specific neural functioning that would correspond to these constructs. Specifically, we found no relationship between learning and connectivity in AN-BP and HC, possibly indicating that these learning systems might not be encoded in the connectivity signatures investigated here but may be more distributed, particularly as our sample is adolescent. Future work should replicate AN-BP findings, examine impact of illness state, and utilize task-based neuroimaging to reveal corresponding neural functioning. Lastly, we recognize that the dual-system framework may have limitations. Collins & Cockburn (2020)^89^ have argued that the forced dichotomy of the model-based/model-free framework artificially limit the dimensionality of learning processes. Moreover, model-based and model-free systems may not be as dissociable as this framework suggests. Consequently, future work, as suggested by Collins & Cockburn (2020), should explore different dimensions of learning systems, as well as the constructs that may influence components of those systems (e.g., uncertainty).

### Conclusions and clinical implications

This is the first study to evaluate the contribution of model-based and model-free learning for reward and punishment in adolescent AN-R and AN-BP. Using a well-validated two-step decision-making task and rsfMRI, we observed important distinctions primarily between AN-R and HC, suggesting increased reliance on model-free learning for reward (e.g., habit) may contribute to the ability to consistently restrict food in AN-R. Exploratory results also suggest that a stronger relationship between model-based learning for reward and neural connectivity in reward circuitry may serve as a protective factor for AN-R. Finally, there is preliminary evidence of distinctions between AN subtypes that may result in divergence of mechanisms contributing to symptom expression. Consequently, interventions that reduce model-free learning and/or improve model-based learning may facilitate recovery in adolescence, though this may be subtype-dependent.

## Data Availability

Data will be available upon reasonable request once the manuscript has been published.

## Acknowledgements and Disclosures

This work was supported by National Institutes of Health Grant No. R21MH118409. C.B. is funded by NIH/NIMH Grant No. F31MH133362. S.D. is funded from the Natural Sciences and Engineering Research Council of Canada.

The authors report no biomedical financial interests or potential conflicts of interest.

## References

1. Harris EC, Barraclough B. Excess mortality of mental disorder. Br J Psychiatry. 1998;173(JULY):11–53. doi:10.1192/bjp.173.1.11

2. Mitchell JE, Crow S. Medical complications of anorexia nervosa and bulimia nervosa. Curr Opin Psychiatry. 2006;19(4). https://journals.lww.com/co-psychiatry/Fulltext/2006/07000/Medical_complications_of_anorexia_nervosa_and.19.aspx

3. Braun DL, Sunday SR, Halmi KA. Psychiatric comorbidity in patients with eating disorders. Psychol Med. 1994;24(4):859–867. doi:DOI: 10.1017/S0033291700028956

4. Arcelus J, Mitchell AJ, Wales J, Nielsen S. Mortality rates in patients with anorexia nervosa and other eating disorders: A meta-analysis of 36 studies. Arch Gen Psychiatry. 2011;68(7):724–731. doi:10.1001/archgenpsychiatry.2011.74

5. Grange D le, Loeb KL. Early identification and treatment of eating disorders: prodrome to syndrome. Early Interv Psychiatry. 2007;1(1):27–39. doi:10.1111/j.1751-7893.2007.00007.x

6. Uniacke B, Timothy Walsh B, Foerde K, Steinglass J. The role of habits in anorexia nervosa: Where we are and where to go from here? Curr Psychiatry Rep. 2018;20(8). doi:10.1007/s11920-018-0928-5

7. Walsh BT. The enigmatic persistence of anorexia nervosa. Am J Psychiatry. 2013;170(5):477–484. doi:10.1176/appi.ajp.2012.12081074

8. Steinglass JE, Walsh BT. Neurobiological model of the persistence of anorexia nervosa. J Eat Disord. 2016;4(1):1–7. doi:10.1186/s40337-016-0106-2

9. Steding J, Boehm I, King JA, et al. Goal-directed vs. habitual instrumental behavior during reward processing in anorexia nervosa: an fMRI study. Sci Rep. 2019;9(1):1–9. doi:10.1038/s41598-019-49884-6

10. Dolan RJ, Dayan P. Goals and habits in the brain. Neuron. 2013;80(2):312–325. doi:10.1016/j.neuron.2013.09.007

11. Daw ND, Niv Y, Dayan P. Uncertainty-based competition between prefrontal and dorsolateral striatal systems for behavioral control. Nat Neurosci. 2005;8(12):1704–1711. doi:10.1038/nn1560

12. Keramati M, Dezfouli A, Piray P. Speed/accuracy trade-off between the habitual and the goal-directed processes. PLoS Comput Biol. 2011;7(5). doi:10.1371/journal.pcbi.1002055

13. Pezzulo G, Rigoli F, Chersi F. The mixed instrumental controller: Using value of information to combine habitual choice and mental simulation. Front Psychol. 2013;4(MAR):1–15. doi:10.3389/fpsyg.2013.00092

14. Daw ND, Gershman SJ, Seymour B, Dayan P, Dolan RJ. Model-based influences on humans’ choices and striatal prediction errors. Neuron. 2011;69(6):1204–1215. doi:10.1016/j.neuron.2011.02.027

15. Miller KJ, Botvinick MM, Brody CD. Value representations in the rodent orbitofrontal cortex drive learning, not choice. Elife. 2022;11:1–27. doi:10.7554/eLife.64575

16. Tolman EC. Cognitive maps in rats and men. Psychol Rev. 1948;55:189–208. doi:10.1037/h0061626

17. Otto AR, Gershman SJ, Markman AB, Daw ND. The curse of planning: Dissecting multiple reinforcement-learning systems by taxing the central executive. Psychol Sci. 2013;24(5):751–761. doi:10.1177/0956797612463080

18. O’Doherty J, Kringelbach ML, Rolls ET, Hornak J, Andrews C. Abstract reward and punishment representations in the human orbitofrontal cortex. Nat Neurosci. 2001;4(1):95–102. doi:10.1038/82959

19. Valentin V V., Dickinson A, O’Doherty JP. Determining the neural substrates of goal-directed learning in the human brain. J Neurosci. 2007;27(15):4019–4026. doi:10.1523/JNEUROSCI.0564-07.2007

20. Stalnaker TA, Cooch NK, Schoenbaum G. What the orbitofrontal cortex does not do. Nat Neurosci. 2015;18(5):620–627. doi:10.1038/nn.3982

21. Wikenheiser AM, Schoenbaum G. Over the river, through the woods: Cognitive maps in the hippocampus and orbitofrontal cortex. Nat Rev Neurosci. 2016;17(8):513–523. doi:10.1038/nrn.2016.56

22. Balleine BW, O’Doherty JP. Human and rodent homologies in action control: Corticostriatal determinants of goal-directed and habitual action. Neuropsychopharmacology. 2010;35(1):48–69. doi:10.1038/npp.2009.131

23. Mcdannald MA, Takahashi YK, Lopatina N, Pietras BW, Jones JL, Schoenbaum G. Model-based learning and the contribution of the orbitofrontal cortex to the model-free world. Eur J Neurosci. 2012;35(7):991–996. doi:10.1111/j.1460-9568.2011.07982.x

24. Morris LS, Kundu P, Dowell N, et al. Fronto-striatal organization: Defining functional and microstructural substrates of behavioural flexibility. Cortex. 2016;74:118–133. doi:10.1016/j.cortex.2015.11.004

25. Voon V, Derbyshire K, Rück C, et al. Disorders of compulsivity: A common bias towards learning habits. Mol Psychiatry. 2015;20(3):345–352. doi:10.1038/mp.2014.44

26. Groenewegen HJ, Wright CI, Beijer AVJ, Voorn P. Convergence and segregation of ventral striatal inputs and outputs. Ann N Y Acad Sci. 1999;877:49–63. doi:10.1111/j.1749-6632.1999.tb09260.x

27. Talmi D, Seymour B, Dayan P, Dolan RJ. Human pavlovian-instrumental transfer. J Neurosci. 2008;28(2):360-368. doi:10.1523/JNEUROSCI.4028-07.2008

28. Pagnoni G, Zink CF, Montague PR, Berns GS. Activity in human ventral striatum locked to errors of reward prediction. Nat Neurosci. 2002;5(2):97–98. doi:10.1038/nn802

29. Van der Meer MAA, Redish AD. Ventral striatum: A critical look at models of learning and evaluation. Curr Opin Neurobiol. 2011;21(3):387–392. doi:10.1016/j.conb.2011.02.011

30. Sutton RS, Barto AG. Reinforcement Learning an Introduction. MIT Press; 1998.

31. Gillan CM, Otto AR, Phelps EA, Daw ND. Model-based learning protects against forming habits. Cogn Affect Behav Neurosci. 2015;15(3):523–536. doi:10.3758/s13415-015-0347-6

32. de Wit S, Watson P, Harsay HA, Cohen MX, van de Vijver I, Ridderinkhof KR. Corticostriatal connectivity underlies individual differences in the balance between habitual and goal-directed action control. J Neurosci. 2012;32(35):12066–12075. doi:10.1523/JNEUROSCI.1088-12.2012

33. Wan Lee S, Shimojo S, O’Doherty JP. Neural computations underlying arbitration between model-based and model-free learning. Neuron. 2014;81(3):687–699. doi:10.1016/j.neuron.2013.11.028

34. Doll BB, Duncan KD, Simon DA, Shohamy D, Daw ND. Model-based choices involve prospective neural activity. Nat Neurosci. 2015;18(5):767–772. doi:10.1038/nn.3981

35. Patterson TK, Knowlton BJ. Subregional specificity in human striatal habit learning: a meta-analytic review of the fMRI literature. Curr Opin Behav Sci. 2018;20:75–82. doi:10.1016/j.cobeha.2017.10.005

36. Nachev P, Kennard C, Husain M. Functional role of the supplementary and pre-supplementary motor areas. Nat Rev Neurosci. 2008;9(11):856–869. doi:10.1038/nrn2478

37. Brown VM, Chen J, Gillan CM, Price RB. Improving the reliability of computational analyses: Model-based planning and its relationship with compulsivity. Biol Psychiatry Cogn Neurosci Neuroimaging. 2020;5(6):601–609. doi:10.1016/j.bpsc.2019.12.019

38. Gillan CM, Kosinski M, Whelan R, Phelps EA, Daw ND. Characterizing a psychiatric symptom dimension related to deficits in goaldirected control. Elife. 2016;5(MARCH2016):1–24. doi:10.7554/eLife.11305

39. Janssen LK, Mahner FP, Schlagenhauf F, Deserno L, Horstmann A. Reliance on model-based and model-free control in obesity. Sci Rep. 2020;10(1):1–14. doi:10.1038/s41598-020-79929-0

40. Voon V, Baek K, Enander J, et al. Motivation and value influences in the relative balance of goal-directed and habitual behaviours in obsessive-compulsive disorder. Transl Psychiatry. 2015;5(September). doi:10.1038/tp.2015.165

41. Foerde K, Daw ND, Rufin T, Walsh BT, Shohamy D, Steinglass JE. Deficient goal-directed control in a population characterized by extreme goal pursuit. J Cogn Neurosci. 2021;33(3):463–481. doi:10.1162/jocn_a_01655

42. Decker JH, Otto AR, Daw ND, Hartley CA. From creatures of habit to goal-directed learners: Tracking the developmental emergence of model-based reinforcement learning. Psychol Sci. 2016;27(6):848–858. doi:10.1177/0956797616639301

43. Eryilmaz H, Rodriguez-Thompson A, Tanner AS, Giegold M, Huntington FC, Roffman JL. Neural determinants of human goal-directed vs. habitual action control and their relation to trait motivation. Sci Rep. 2017;7(1):1–11. doi:10.1038/s41598-017-06284-y

44. Voon V, Baek K, Enander J, et al. Motivation and value influences in the relative balance of goal-directed and habitual behaviours in obsessive-compulsive disorder. Transl Psychiatry. 2015;5(August). doi:10.1038/tp.2015.165

45. Brooks SJ, Rask-Andersen M, Benedict C, Schiöth HB. A debate on current eating disorder diagnoses in light of neurobiological findings: is it time for a spectrum model? BMC Psychiatry. 2012;12. doi:10.1186/1471-244X-12-76

46. Fladung AK, Schulze UME, Schöll F, Bauer K, Grön G. Role of the ventral striatum in developing anorexia nervosa. Transl Psychiatry. 2013;3(July):2–7. doi:10.1038/tp.2013.88

47. Keating C, Tilbrook AJ, Rossell SL, Enticott PG, Fitzgerald PB. Reward processing in anorexia nervosa. Neuropsychologia. 2012;50(5):567–575. doi:10.1016/j.neuropsychologia.2012.01.036

48. Monteleone AM, Monteleone P, Esposito F, et al. Altered processing of rewarding and aversive basic taste stimuli in symptomatic women with anorexia nervosa and bulimia nervosa: An fMRI study. J Psychiatr Res. 2017;90:94–101. doi:10.1016/j.jpsychires.2017.02.013

49. O’Hara CB, Campbell IC, Schmidt U. A reward-centred model of anorexia nervosa: A focussed narrative review of the neurological and psychophysiological literature. Neurosci Biobehav Rev. 2015;52:131–152. doi:10.1016/j.neubiorev.2015.02.012

50. Wu M, Brockmeyer T, Hartmann M, Skunde M, Herzog W, Friederich HC. Reward-related decision making in eating and weight disorders: A systematic review and meta-analysis of the evidence from neuropsychological studies. Neurosci Biobehav Rev. 2016;61:177–196. doi:10.1016/j.neubiorev.2015.11.017

51. Bischoff-Grethe A, McCurdy D, Grenesko-Stevens E, et al. Altered brain response to reward and punishment in adolescents with Anorexia nervosa. Psychiatry Res - Neuroimaging. 2013;214(3):331–340. doi:10.1016/j.pscychresns.2013.07.004

52. Glashouwer KA, Bloot L, Veenstra EM, Franken IHA, de Jong PJ. Heightened sensitivity to punishment and reward in anorexia nervosa. Appetite. 2014;75:97–102. doi:10.1016/j.appet.2013.12.019

53. Harrison A, O’Brien N, Lopez C, Treasure J. Sensitivity to reward and punishment in eating disorders. Psychiatry Res. 2010;177(1-2):1–11. doi:10.1016/j.psychres.2009.06.010

54. Jappe LM, Frank GKW, Shott ME, et al. Heightened sensitivity to reward and punishment in anorexia nervosa. Int J Eat Disord. 2011;44(4):317–324. doi:10.1002/eat.20815

55. Matton A, de Jong P, Goossens L, et al. Sensitivity for cues predicting reward and punishment in young women with eating disorders. Eur Eat Disord Rev. 2017;25(6):501–511. doi:10.1002/erv.2541

56. Bernardoni F, Geisler D, King JA, et al. Altered medial frontal feedback learning signals in anorexia nervosa. Biol Psychiatry. 2018;83(3):235–243. doi:10.1016/j.biopsych.2017.07.024

57. Wierenga CE, Reilly E, Bischoff-Grethe A, Kaye WH, Brown GG. Altered reinforcement learning from reward and punishment in anorexia nervosa: Evidence from computational modeling. J Int Neuropsychol Soc. 2021;(2021):1–13. doi:10.1017/S1355617721001326

58. O’hara CB, Campbell IC, Schmidt U. A reward-centred model of anorexia nervosa: A focussed narrative review of the neurological and psychophysiological literature. Neurosci Biobehav Rev. 2015;52:131–152. doi:10.1016/j.neubiorev.2015.02.012

59. Wierenga CE, Ely A, Bischoff-Grethe A, Bailer UF, Simmons AN, Kaye WH. Are extremes of consumption in eating disorders related to an altered balance between reward and inhibition? Front Behav Neurosci. 2014;8(DEC):410. doi:10.3389/fnbeh.2014.00410

60. Reilly EE, Rockwell RE, Ramirez AL, et al. Naturalistic outcomes for a day-hospital programme in a mixed diagnostic sample of adolescents with eating disorders. Eur Eat Disord Rev. 2020;28(2):199–210. doi:10.1002/erv.2716

61. Otto AR, Raio CM, Chiang A, Phelps EA, Daw ND. Working-memory capacity protects model-based learning from stress. Proc Natl Acad Sci U S A. 2013;110(52):20941–20946. doi:10.1073/pnas.1312011110

62. Bates D, Mächler M, Bolker BM, Walker SC. Fitting linear mixed-effects models using lme4. J Stat Softw. 2015;67(1). doi:10.18637/jss.v067.i01

63. R Development Core Team. R: A language and environment for statistical computing. In: R Foundation for Statistical Computing.; 2015. doi:10.1007/978-1-4020-6850-8

64. Marcus DS, Harms MP, Snyder AZ, et al. Human Connectome Project informatics: Quality control, database services, and data visualization. Neuroimage. 2013;80:202–219. doi:10.1016/j.neuroimage.2013.05.077

65. Glasser MF, Sotiropoulos SN, Wilson JA, et al. The Minimal preprocessing pipelines for the Human Connectome Project and for the WU-Minn HCP Consortium. Neuroimage. 2013;80:105–12404. doi:10.1016/j.neuroimage.2013.04.127.The

66. Beckmann CF, Smith SM. Probabilistic independent component analysis for functional magnetic resonance imaging. IEEE Trans Med Imaging. 2004;23(2):137–152. doi:10.1109/TMI.2003.822821

67. Glasser MF, Smith SM, Marcus DS, et al. The Human Connectome Project’s neuroimaging approach. Nat Neurosci. 2016;19(9):1175–1187. doi:10.1038/nn.4361

68. Desikan RS, Ségonne F, Fischl B, et al. An automated labeling system for subdividing the human cerebral cortex on MRI scans into gyral based regions of interest. Neuroimage. 2006;31(3):968–980. doi:10.1016/j.neuroimage.2006.01.021

69. Frazier JA, Chiu S, Breeze JL, et al. Structural brain magnetic resonance imaging of limbic and thalamic volumes in pediatric bipolar disorder. Am J Psychiatry. 2005;162(7):1256–1265. doi:10.1176/appi.ajp.162.7.1256

70. Makris N, Goldstein JM, Kennedy D, et al. Decreased volume of left and total anterior insular lobule in schizophrenia. Schizophr Res. 2006;83(2):155–171. 10.1016/j.schres.2005.11.020

71. Goldstein JM, Seidman LJ, Makris N, et al. Hypothalamic abnormalities in schizophrenia: Sex effects and genetic vulnerability. Biol Psychiatry. 2007;61(8):935–945. 10.1016/j.biopsych.2006.06.027

72. Eppinger B, Walter M, Li SC. Electrophysiological correlates reflect the integration of model-based and model-free decision information. Cogn Affect Behav Neurosci. 2017;17(2):406–421. doi:10.3758/s13415-016-0487-3

73. Steinglass J, Walsh BT. Habit learning and anorexia nervosa: A cognitive neuroscience hypothesis. Int J Eat Disord. 2006;39(4):267–275. doi:10.1002/eat.20244

74. Godier LR, de Wit S, Pinto A, et al. An investigation of habit learning in Anorexia Nervosa. Psychiatry Res. 2016;244:214–222. doi:10.1016/j.psychres.2016.07.051

75. Haynos AF, Hall LMJ, Lavender JM, et al. Resting state functional connectivity of networks associated with reward and habit in anorexia nervosa. Hum Brain Mapp. 2019;40(2):652–662. doi:10.1002/hbm.24402

76. Cha J, Ide JS, Bowman FD, Simpson HB, Posner J, Steinglass JE. Abnormal reward circuitry in anorexia nervosa: A longitudinal, multimodal MRI study. Hum Brain Mapp. 2016;37(11):3835–3846. doi:10.1002/hbm.23279

77. Kaye WH, Wierenga CE, Bailer UF, Simmons AN, Bischoff-Grethe A. Nothing tastes as good as skinny feels: The neurobiology of anorexia nervosa. Trends Neurosci. 2013;36(2):110–120. doi:10.1016/j.tins.2013.01.003

78. Wierenga CE, Ely A, Bischoff-Grethe A, Bailer UF, Simmons AN, Kaye WH. Are extremes of consumption in eating disorders related to an altered balance between reward and inhibition? Front Behav Neurosci. 2014;8(DEC). doi:10.3389/fnbeh.2014.00410

79. Dang J, King KM, Inzlicht M. Why are self-report and behavioral measures weakly correlated? Trends Cogn Sci. 2020;24(4):267–269. doi:10.1016/j.tics.2020.01.007

80. Bolenz F, Eppinger B. Valence bias in metacontrol of decision making in adolescents and young adults. Child Dev. 2022;93(2):e103–e116. doi:10.1111/cdev.13693

81. Scholz V, Waltmann M, Herzog N, Reiter A, Horstmann A, Deserno L. Cortical grey matter mediatesincreases in model-based control and learning from positive feedback from adolescence to adulthood. J Neurosci. 2023;43(12):JN-RM-1418–22. doi:10.1523/jneurosci.1418-22.2023

82. Vaghi MM, Moutoussis M, Váša F, et al. Compulsivity is linked to reduced adolescent development of goal-directed control and frontostriatal functional connectivity. Proc Natl Acad Sci U S A. 2020;117(41):25911–25922. doi:10.1073/pnas.1922273117

83. Potter TCS, Bryce N V., Hartley CA. Cognitive components underpinning the development of model-based learning. Dev Cogn Neurosci. 2017;25:272–280. doi:10.1016/j.dcn.2016.10.005

84. Beck I, Smits DJM, Claes L, Vandereycken W, Bijttebier P. Psychometric evaluation of the behavioral inhibition/behavioral activation system scales and the sensitivity to punishment and sensitivity to reward questionnaire in a sample of eating disordered patients. Pers Individ Dif. 2009;47(5):407–412. doi:10.1016/j.paid.2009.04.007

85. Claes L, Nederkoorn C, Vandereycken W, Guerrieri R, Vertommen H. Impulsiveness and lack of inhibitory control in eating disorders. Eat Behav. 2006;7(3):196–203. doi:10.1016/j.eatbeh.2006.05.001

86. Keel PK, Kennedy GA, Rogers ML, et al. Reliability and validity of a transdiagnostic measure of reward valuation effort. Psychol Assess. 2022;34(5):419–430. doi:10.1037/pas0001107

87. Otto AR, Skatova A, Madlon-Kay S, Daw ND. Cognitive control predicts use of model-based reinforcement learning. J Cogn Neurosci. 2015;27(2):319–333. doi:10.1162/jocn_a_00709

88. Worbe Y, Palminteri S, Savulich G, et al. Valence-dependent influence of serotonin depletion on model-based choice strategy. Mol Psychiatry. 2016;21(5):624–629. doi:10.1038/mp.2015.46

89. Collins AGE, Cockburn J. Beyond dichotomies in reinforcement learning. Nat Rev Neurosci. 2020;21(10):576–586. doi:10.1038/s41583-020-0355-6

