## Supplement for "Greater reliance on model-free learning in adolescent anorexia nervosa: An examination of dual-system reinforcement learning"

**Habit and Goal-Directed Learning in Adolescent Anorexia Nervosa**

***Participants***

*Inclusion/Exclusion Criteria*

To participate in the study, AN must: (a) have met DSM-5 criteria for AN-R or AN-BP, (b) be medically stable, per American Academy of Pediatrics and the Society of Adolescent Medicine requirements; and (c) be at least 75% of adjusted ideal body weight (IBW). HCs were recruited from the San Diego community and were administered the Kiddie Schedule for Affective Disorders and Schizophrenia for School‐Age Children (KSADS; Kaufman et al., 1997) and the Eating Disorder Examination (EDE; Cooper & Fairburn, 1987) to confirm the absence of symptoms related to any eating disorder or Axis I psychiatric disorder. HCs were included in the study if they had: (a) no stigmata suggestive of an eating disorder, (b) no current or past psychiatric (definitive Axis I disorder) or medical illness; (c) no use of psychoactive drugs; and (d) maintained 90% to 120% ideal body weight since menarche. Exclusion criteria for all subjects included the following: (a) MRI contraindications, (b) psychotic illness/other mental illness requiring hospitalization; (c) current dependence on drugs or alcohol; (d) physical conditions (e.g., diabetes mellitus, pregnancy) known to influence eating or weight; (e) neurological disorder or history of head injury with >30 min loss of consciousness; (f) neurodevelopmental disorder; (g) primary obsessive compulsive disorder or primary major depressive disorder (within last 3 months as measured by the KSADS).

***Self-Report Measures***

The State-Trait Anxiety Inventory – Trait subscale (STAI-T; Spielberger, Gorsuch, & Lushene, 1970) is a 20-item self-report measure that assesses trait anxiety on a 1- 4 Likert scale, with higher values indicating greater severity of symptoms.

The Beck Depression Inventory – II (BDI-II; Beck et al., 1996) is a 21-item self-report measure assessing severity of depression symptoms. Items are rated on a 0 – 3 Likert scale.

The Temperament and Character Inventory (TCI; Cloninger et al., 1994) is a 226-item self-report measure assessing seven dimensions of personality. For the purposes of the current study, we only include the Harm Avoidance subscale, designed to correspond with the magnitude of inhibitory responding towards aversive stimuli.

Sensitivity to Punishment and Reward Questionnaire (SPSRQ; Torrubia et al., 2001) is a 48-item self-report measure, including Punishment and Reward subscales, designed to operationalize constructs from reinforcement learning theory. We included both subscale scores in our analyses. Items are rated as binary (yes/no) responses.

Behavioral Inhibition/Behavioral Activation Scales (BIS/BAS; Carver & White, 1994) is a 24-item self-report measure, which includes a Behavioral Inhibition (BIS) scale and three intercorrelated Behavioral Activation (BAS) subscales: Reward Responsiveness, Drive, and Fun Seeking. We only included BIS and BAS-Reward Responsiveness in our analyses.

Adult Temperament Questionnaire (ATQ; Evans & Rothbart, 2007) is a 77-item self-report questionnaire measuring temperamental constructs, including Effortful Control, Negative Affect, Extraversion, and Orienting Sensitivity. We only included the Effortful Control subscale in our analyses, reflecting self-regulation and executive control.

***Two-Step Sequential Decision-Making Task***

Participants were tasked with collecting “space treasure” that would amount to a $1 gain in the reward condition or no monetary loss (of $1) in the punishment condition. On each trial, participants made choices at two stages. In the first stage they made a choice between two spaceships that traveled to one of two planets. Each spaceship traveled more frequently to one planet (70%) and less frequently to the other (30%); for example, the blue rocketship had a 70% chance of traveling to the red planet (a common transition) and a 30% chance of traveling to the purple planet (a rare transition), while the green rocketship had the opposite set of probabilities. At the second stage (i.e., when participants are on a planet), they were asked to choose between two aliens. Depending on which alien they chose, they were either presented with space treasure (+$1) or with an empty circle for the reward condition. For the punishment condition, they were either presented with an empty circle or space treasure with a red ‘x’ over it (-$1). Probability of winning any given trial was determined by a slowly drifting probability bounded between 0.2 and 0.8. During the task, participants were allotted three seconds to make a decision, followed by a one-second animation, a one-second feedback period, and a one-second inter-trial interval. The game itself consisted of four blocks separated by breaks and comprised of 200 trials.

***Computational Model***

The hybrid reinforcement learning model (Decker et al., 2016) is comprised of two distinct components: a model-free SARSA (𝜆) temporal difference algorithm and a model-based “tree-search” reinforcement learning algorithm (Bellman’s equation), enabling representation of all possible choice options and associated outcomes (Sutton & Barto, 1998). The task defines three states ($i$): one state at stage one ($s_{A}$) and two states at stage two ($s_{B}, s_{C}$), where each state is associated with a set of actions ($a_{A}, a_{B}$). Both algorithms track state-action value functions, Q(*s,a*), that represent the expected value for each state-action pair. For every trial, $t$, the participant makes a stage 1 choice ($a_{1, i,t}$), followed by a transition to the second stage where they again make a choice ($a_{i,2, t}$), that results in a possible reward ($r_{i,t}$).

*Model-free algorithm*

The model-free temporal difference algorithm updates the state action values according to the following rule:

$$Q_{MF}\left( s_{i, t}, a_{i, t} \right)= Q_{MF}\left( s_{i, t-1}, a_{i, t-1} \right)+ \alpha\delta_{i, t}$$

Where, $\delta_{i, t}= r_{i, t}+ Q_{MF}\left( s_{i+1, t}, a_{i+1,t} \right)- Q_{MF}\left( s_{i, t-1}, a_{i, t-1} \right)$

Here, the reward prediction error (RPE) is defined by $\delta$, and the learning rate is defined by $\alpha$. For stage one, $r_{1,t}=0$ and the RPE is calculated with respect to the estimate of the second stage action value, $Q_{MF}\left( s_{2, t}, a_{2,t} \right)$. For the second stage, $r_{2,t}=0$, for a loss, $r_{2,t}=1$, for a reward, or $r_{2,t}= -1$ for a punishment and the delta rule simplifies to $\delta_{i, t}= r_{i, t}- Q_{MF}\left( s_{2, t-1}, a_{2, t-1} \right)$.

*Model-based algorithm*

The model-based algorithm was applied only at the first stage and updates the state-action pair by incorporating information about the 70/30 transition probability structure:

$$Q_{MB}\left( s_{1, t}, a_{j, t} \right)=P\left( s_{B} | s_{A}, a_{j} \right) \max_{a\epsilon\left\{ a_{A}, a_{B} \right\}} Q_{TD}\left( S_{B},a \right)+ P\left( s_{C} | s_{A}, a_{j} \right) \max_{a\epsilon\left\{ a_{A}, a_{B} \right\}} Q_{TD}\left( S_{C},a \right)$$

*Choice rule*

We applied a softmax choice rule as our policy, calculating a probability to each action according to the combination of both $Q_{MB}$ and $Q_{MF}$ values taken from the model-based and model-free algorithms described above. Each value was weighted with its own inverse temperature parameter, $\beta_{MB}$ and $\beta_{MF}$, to ascertain the relative contribution of model-based and model-free valuations:

$$P(a_{i,t}=a|s_{i,t})= \frac{exp\left[ \beta_{MF}* Q_{MF}\left( s_{i,t}, a \right)+ \beta_{MB}* Q_{MB}\left( s_{i,t},a \right)+p*rep(a) \right]}{\sum a^{'}exp\left[ \beta_{MF}* Q_{MF}\left( s_{i,t}, a \right)+ \beta_{MB}* Q_{MB}\left( s_{i,t},a \right)+p*rep(a^{'}) \right]}$$

Here, rep(a) is an indicator function that takes on a value of 1 for a first-stage action that repeats the action taken in the previous trial. The "stickiness" parameter (𝑝) is applied to measure perseveration (𝑝>0) or switching (𝑝<0) behavior by combining it with the indicator function. Model-based learning is only possible at the first stage, so the softmax function simplifies to only include $Q_{MF}$ with its own inverse temperature, $\beta_{2}.$

*Group level modeling*

The equations above represent modelling for a single subject, $j$, which was embedded within a multi-level random effects model. The free parameters of the model (α, $\beta_{MB}$, $\beta_{MF}$, $\beta_{2}$, 𝑝) were taken as random effects for each subject, *j,* and extracted from a common group level distribution.

To test the dependence of the free parameters, $p$, on diagnostic group, diagnostic group was entered into a regression at the group level with infinite support:

$$\mu_{p,j} \sim\gamma_{0}+\gamma_{ANR}is\_R_{j}+\gamma_{ANBP}is\_BP_{j}$$

where $is\_R$ and $is\_BP$ are dummy-coded variables reflecting whether participant $j$ was (or was not) in the AN-R or AN-BP subgroup respectively. To reduce estimation issues arising from the hierarchical nature of the data, free parameters were reparametrized. Specifically, free parameters were initially drawn from a standard normal distribution ($\mu^{UT}$, UT = untransformed) and then transformed into an appropriate scale based on estimated fixed effects ($\gamma$) and independently estimated random variance terms ($\sigma$):

$$\beta_{p,j}=\mu_{p,j}+\sigma_{p,j}\mu_{p,j}^{UT}$$

These parameters were further transformed to enforce appropriate bounds. Namely, inverse temperature parameter was exponentiated (to enforce [0,$\infty$] bounds), learning rates were converted to a unit scale, and the stickiness parameter was converted to a unit scale and multiplied by an upper bound of 5. Hyperparameters $\gamma$ and $\sigma$ were given normal priors of $N(0,1)$ and $HN\left( 0,.2 \right)$ respectively.

*Estimation*

The joint distribution of model parameters was estimated separately for reward and punishment conditions. Markov Chain Monte Carlo (MCMC) techniques, specifically the No-U-Turn variant of Hamiltonian Monte Carlo, implemented in the Stan modeling language (Stan Development Team, 2015), was implemented to obtain samples from the conditional joint distribution. We ran four chains of 4,000 samples each, with the first 1,000 samples of each chain being discarded as burn-in, and no thinning applied.

| Table S1.  *Diagnostic Information* | | | |  |  |
| --- | --- | --- | --- | --- | --- |
| **Variable** | **AN-R (n = 36) ------------**  ***n* (%)** | **AN-BP (n = 20) ------------**  ***n* (%)** | **HC (n = 28) ------------**  ***n* (%)** | ***X^2^*** | ***p*** |
| Lifetime | | | | | |
| Major Depressive Disorder | 19 (52.78) | 13 (65.00) | 0 (0.00) | 26.66 | <.001 |
| Persistent Depressive Disorder | 8 (22.22) | 7 (35.00) | 0 (0.00) | 10.56 | .005 |
| Other Specified Depressive Disorder | 1 (2.78) | 0 (0.00) | 0 (0.00) | 1.39 | .50 |
| Panic Disorder | 0 (0.00) | 1 (5.00) | 0 (0.00) | 3.19 | .20 |
| Social Anxiety Disorder | 12 (33.33) | 6 (30.00) | 0 (0.00) | 11.54 | .003 |
| Specific Phobia | 1 (2.78) | 0 (0.00) | 0 (0.00) | 1.35 | .51 |
| Generalized Anxiety Disorder | 12 (33.33) | 7 (35.00) | 0 (0.00) | 12.30 | .002 |
| Obsessive-Compulsive Disorder | 7 (19.44) | 4 (20.00) | 0 (0.00) | 6.33 | .04 |
| Post-Traumatic Stress Disorder | 1 (2.78) | 1 (5.00) | 0 (0.00) | 1.30 | .52 |
| Attention-Deficit Hyperactivity Disorder | 0 (0.00) | 2 (10.00) | 0 (0.00) | 6.56 | .04 |
| Substance Use Disorder | 0 (0.00) | 1 (5.00) | 0 (0.00) | 3.24 | .20 |
| Current | | | | | |
| Major Depressive Disorder | 15 (41.67) | 12 (60.00) | 0 (0.00) | 21.88 | <.001 |
| Persistent Depressive Disorder | 7 (19.44) | 7 (35.00) | 0 (0.00) | 10.64 | .005 |
| Other Specified Depressive Disorder | 1 (2.78) | 0 (0.00) | 0 (0.00) | 1.32 | .52 |
| Panic Disorder | 0 (0.00) | 1 (5.00) | 0 (0.00) | 3.24 | .20 |
| Social Anxiety Disorder | 12 (33.33) | 5 (25.00) | 0 (0.00) | 11.21 | .004 |
| Specific Phobia | 1 (2.78) | 0 (0.00) | 0 (0.00) | 1.35 | .51 |
| Generalized Anxiety Disorder | 11 (30.56) | 7 (35.00) | 0 (0.00) | 11.61 | .003 |
| Obsessive-Compulsive Disorder | 7 (19.44) | 3 (15.00) | 0 (0.00) | 5.92 | .05 |
| Post-Traumatic Stress Disorder | 1 (2.78) | 0 (0.00) | 0 (0.00) | 1.35 | .52 |
| Attention-Deficit Hyperactivity Disorder | 0 (0.00) | 2 (10.00) | 0 (0.00) | 6.56 | .04 |
| Substance Use Disorder | 0 (0.00) | 1 (5.00) | 0 (0.00) | 3.24 | .20 |

*Note*. AN-R – restricting type anorexia nervosa; AN-BP – binge/purge type anorexia nervosa; HC – healthy controls

| Table S2.  *Raw Choice Data Mixed Effects Logistic Regression (4-way interaction)* | | | | |  |
| --- | --- | --- | --- | --- | --- |
| **Variable** | **Odds Ratio** | **Standard Error** | | **Statistic** | ***p*** |
| Intercept [AN-R] | 0.81 | | 0.14 | 5.83 | **<.001** |
| Last won | 0.95 | | 0.2 | 4.83 | **<.001** |
| Last transition | 0.22 | | 0.13 | 1.75 | .08 |
| Condition | -0.16 | | 0.1 | -1.67 | .10 |
| AN-BP | 0.15 | | 0.23 | 0.65 | .52 |
| HC | -0.21 | | 0.21 | -1.03 | .30 |
| Last won x last transition | -0.71 | | 0.2 | -3.46 | **<.001** |
| Last won x condition | 0.35 | | 0.17 | 2.03 | **.04** |
| Last transition x condition | 0.21 | | 0.13 | 1.62 | .10 |
| Last won x AN-BP | 0.33 | | 0.33 | 0.98 | .33 |
| Last won x HC | 0.16 | | 0.3 | 0.54 | .59 |
| Last transition x AN-BP | -0.04 | | 0.21 | -0.21 | .83 |
| Last transition x HC | 0.18 | | 0.19 | 0.94 | .35 |
| Condition x AN-BP | -0.09 | | 0.16 | -0.56 | .57 |
| Condition x HC | 0.12 | | 0.15 | 0.81 | .42 |
| Last won x last transition x condition | -0.28 | | 0.26 | -1.08 | .28 |
| Last won x last transition x AN-BP | -0.05 | | 0.35 | -0.16 | .88 |
| Last won x last transition x HC | 0.02 | | 0.31 | 0.08 | .94 |
| Last won x condition x AN-BP | -0.23 | | 0.29 | -0.8 | .42 |
| Last won x condition x HC | -0.23 | | 0.26 | -0.9 | .37 |
| Last transition x condition x AN-BP | -0.04 | | 0.22 | -0.17 | .87 |
| Last transition x condition x HC | -0.06 | | 0.2 | -0.29 | .78 |
| Last won x last transition x condition x AN-BP | -0.4 | | 0.43 | -0.93 | .35 |
| Last won x last transition x condition x HC | -0.18 | | 0.39 | -0.46 | .64 |

*Note*. AN-R – restricting type anorexia nervosa; AN-BP – binge/purge type anorexia nervosa; HC – healthy controls

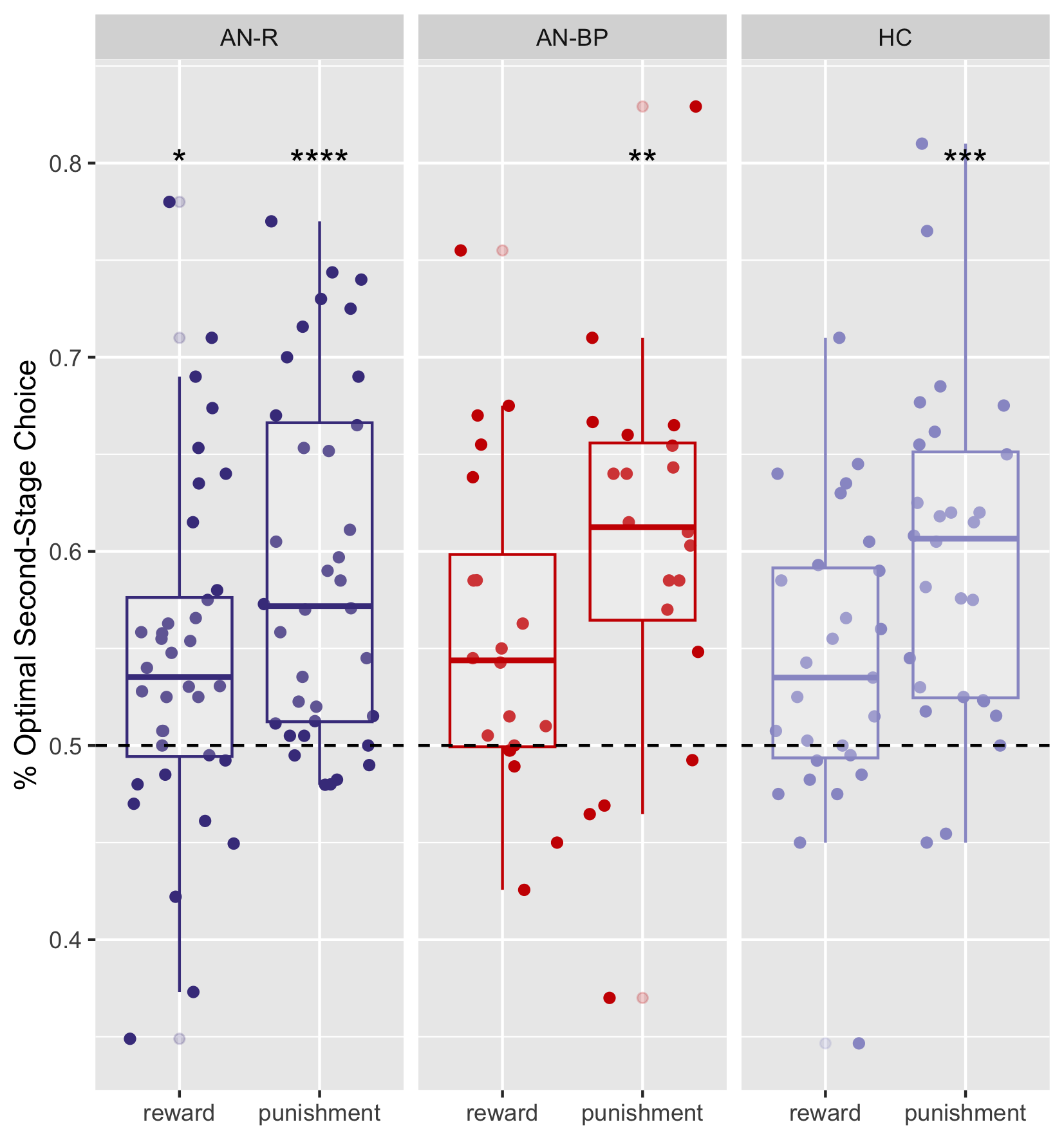

**Figure S1**. Percentage of second-stage optimal choice plots for each group and each condition. The dashed line is at chance, with mean values for each group and condition compared to chance in one-way Wilcox tests. * <.05, **<.01, ***<.001

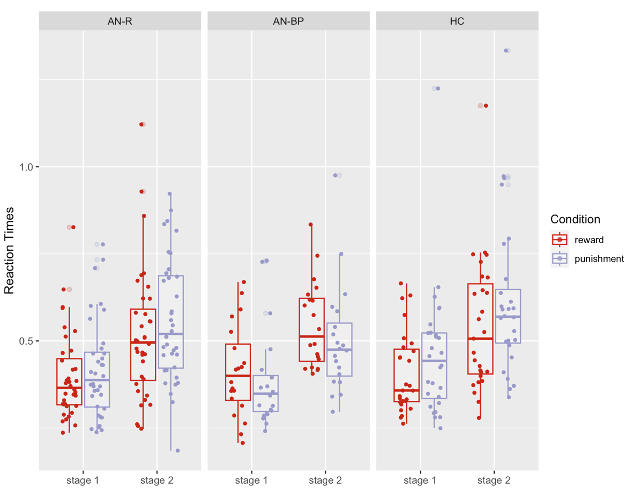

**Figure S2**. Reaction time data during first and second stage choices, for each group and condition. There were no significant group differences, nor were there differences by condition.

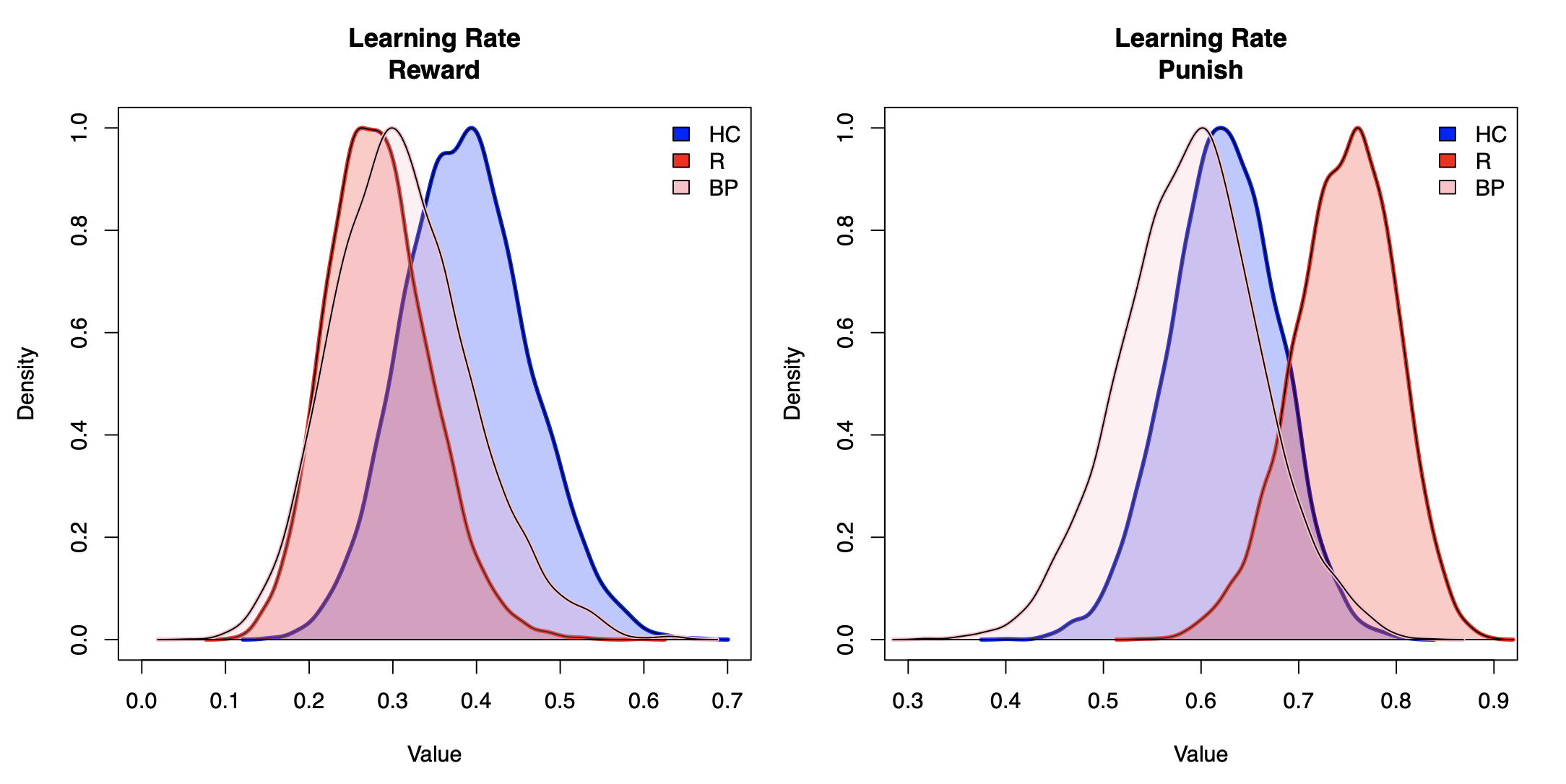

**Figure S3**. Group-level posterior distributions of learning rates ($\gamma_{0}$, $\gamma_{1}$, $\gamma_{2}$) for reward (left) and punishment (right). Values have been transformed using the inverse cumulative distribution function to display estimates on the correct scale. R – restricting type anorexia nervosa, BP – binge/purge type anorexia nervosa, HC – healthy control

| Table S3.  *Latent Regression Coefficients on Learning Rate* | | | |  |
| --- | --- | --- | --- | --- |
| **Parameter** | **Mean Posterior** | ***p*** | **CI[2.5%]** | **CI[97.5%]** |
| *Reward* | | | | |
| $\gamma_{0}$ [HC] | -0.29 | .07 | -0.68 | -0.42 |
| $\gamma_{1}$ [AN-R vs HC] | -0.29 | .14 | -0.81 | 0.23 |
| $\gamma_{2}$ [AN-BP vs HC] | -0.21 | .24 | -0.81 | 0.37 |
| *Punishment* | | | | |
| $\gamma_{0}$ [HC] | 0.32 | .02 | 0.03 | 0.61 |
| $\gamma_{1}$ [AN-R vs HC] | 0.35 | .05 | -0.08 | 0.79 |
| $\gamma_{2}$ [AN-BP vs HC] | -0.09 | .36 | -0.55 | 0.39 |

*Note*. AN-R – restricting type anorexia nervosa; AN-BP – binge/purge type anorexia nervosa; HC – healthy controls

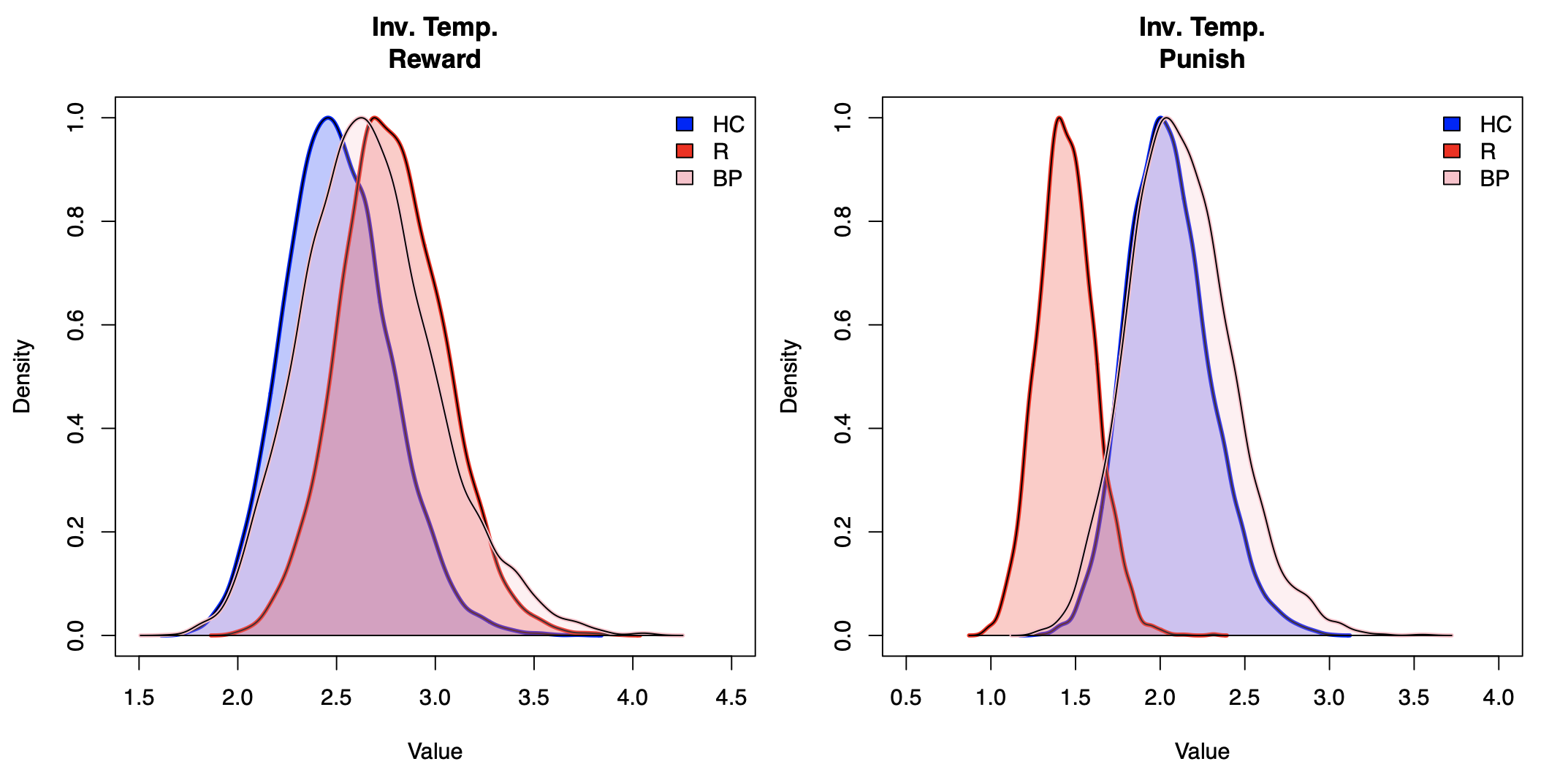

**Figure S4**. Group-level posterior distributions of second-stage inverse temperature parameters ($\gamma_{0}$, $\gamma_{1}$, $\gamma_{2}$) for reward (left) and punishment (right). Values have been log transformed to display estimates on the correct scale. R – restricting type anorexia nervosa, BP – binge/purge type anorexia nervosa, HC – healthy control

| Table S4.  *Latent Regression Coefficients on Inverse Temperature* | | | |  |
| --- | --- | --- | --- | --- |
| **Parameter** | **Mean Posterior** | ***p*** | **CI[2.5%]** | **CI[97.5%]** |
| *Reward* | | | | |
| $\gamma_{0}$ [HC] | 0.91 | 0 | 0.71 | 1.11 |
| $\gamma_{1}$ [AN-R vs HC] | 0.11 | .22 | 0.17 | 0.38 |
| $\gamma_{2}$ [AN-BP vs HC] | 0.06 | .36 | -0.26 | 0.38 |
| *Punishment* | | | | |
| $\gamma_{0}$ [HC] | 0.71 | 0 | 0.48 | 0.94 |
| $\gamma_{1}$ [AN-R vs HC] | -0.35 | .01 | -0.67 | -0.04 |
| $\gamma_{2}$ [AN-BP vs HC] | 0.03 | .43 | -0.33 | 0.37 |

*Note*. AN-R – restricting type anorexia nervosa; AN-BP – binge/purge type anorexia nervosa; HC – healthy controls

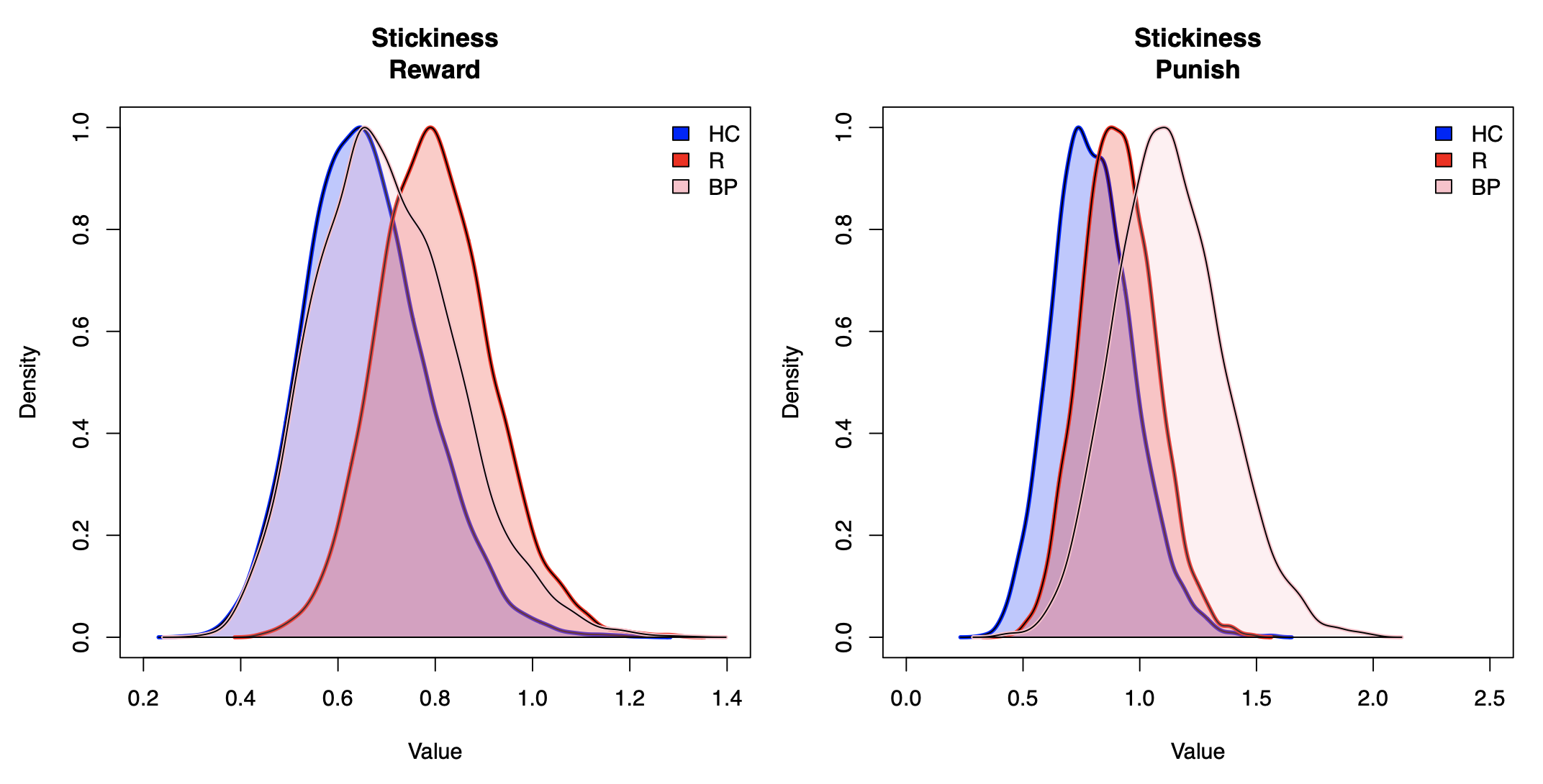

**Figure S5**. Group-level posterior distributions of stickiness parameters ($\gamma_{0}$, $\gamma_{1}$, $\gamma_{2}$) for reward (left) and punishment (right). Values have been log transformed using the inverse cumulative distribution function to display estimates on the correct scale. R – restricting type anorexia nervosa, BP – binge/purge type anorexia nervosa, HC – healthy control

| Table S5.  *Latent Regression Coefficients on Stickiness Parameter* | | | |  |
| --- | --- | --- | --- | --- |
| **Parameter** | **Mean Posterior** | ***p*** | **CI[2.5%]** | **CI[97.5%]** |
| *Reward* | | | | |
| $\gamma_{0}$ [HC] | -1.13 | 0 | -1.35 | -0.90 |
| $\gamma_{1}$ [AN-R vs HC] | 0.13 | .20 | -0.16 | 0.42 |
| $\gamma_{2}$ [AN-BP vs HC] | 0.04 | .42 | -0.29 | 0.37 |
| *Punishment* | | | | |
| $\gamma_{0}$ [HC] | -1.00 | <.001 | -1.28 | -0.73 |
| $\gamma_{1}$ [AN-R vs HC] | 0.09 | .32 | -0.28 | 0.44 |
| $\gamma_{2}$ [AN-BP vs HC] | 0.24 | .13 | -0.15 | 0.63 |

*Note*. AN-R – restricting type anorexia nervosa; AN-BP – binge/purge type anorexia nervosa; HC – healthy controls

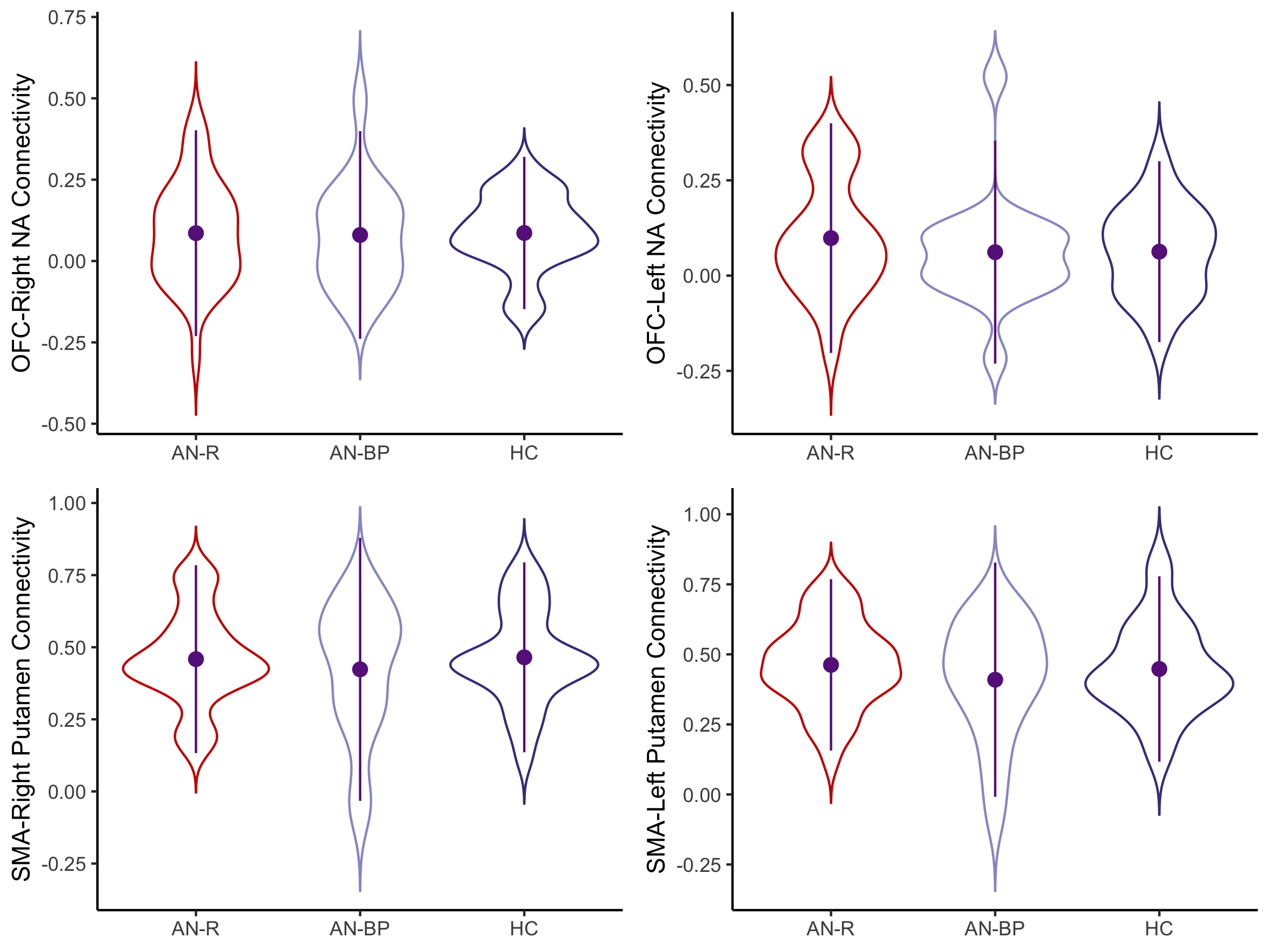

**Figure S6**. Violin plots of correlation coefficients for each ROI-to-ROI dyad in each group. There were no significant group differences in strength of connectivity. AN-R – restricting type anorexia nervosa, AN-BP – binge/purge type anorexia nervosa, HC – healthy control

| Table S6.  *Model-based Weights and OFC-to-left NAcc Mixed Effects Regression* | | | |  |
| --- | --- | --- | --- | --- |
| **Variable** | **Estimate** | **Standard Error** | **Statistic** | ***p*** |
| Intercept [AN-R] | -0.86 | 0.17 | -5.14 | **<.001** |
| AN-BP | 1.51 | 0.27 | 5.56 | **<.001** |
| HC | 1.38 | 0.25 | 5.57 | **<.001** |
| Condition [punishment] | 0.17 | 0.23 | 0.72 | .47 |
| OFC-NAcc connectivity | 2.15 | 0.92 | 2.32 | **.02** |
| Age | 0.00 | 0.07 | 0.07 | .95 |
| BMI | -0.05 | 0.10 | -0.44 | .66 |
| WASI Full Scale IQ | 0.19 | 0.07 | 2.73 | **.007** |
| AN-BP x Condition [punishment] | -0.51 | 0.38 | -1.34 | .18 |
| HC x Condition [punishment] | -0.15 | 0.34 | -0.43 | .67 |
| AN-BP x OFC-NAcc connectivity | -2.27 | 1.67 | -1.36 | .18 |
| HC x OFC-NAcc connectivity | -2.22 | 1.64 | -1.35 | .18 |
| Condition [punishment] x OFC-NAcc connectivity | -1.31 | 1.29 | -1.01 | .32 |
| AN-BP x Condition [punishment] x OFC-NAcc connectivity | -0.17 | 2.32 | -0.07 | .94 |
| HC x Condition [punishment] x OFC-NAcc connectivity | 3.34 | 2.29 | 1.46 | .15 |

*Note*. AN-R – restricting type anorexia nervosa; AN-BP – binge/purge type anorexia nervosa; HC – healthy controls; OFC – orbitofrontal cortex; NAcc – nucleus accumbens; WASI – Weschler Abbreviated Scale of Intelligence.

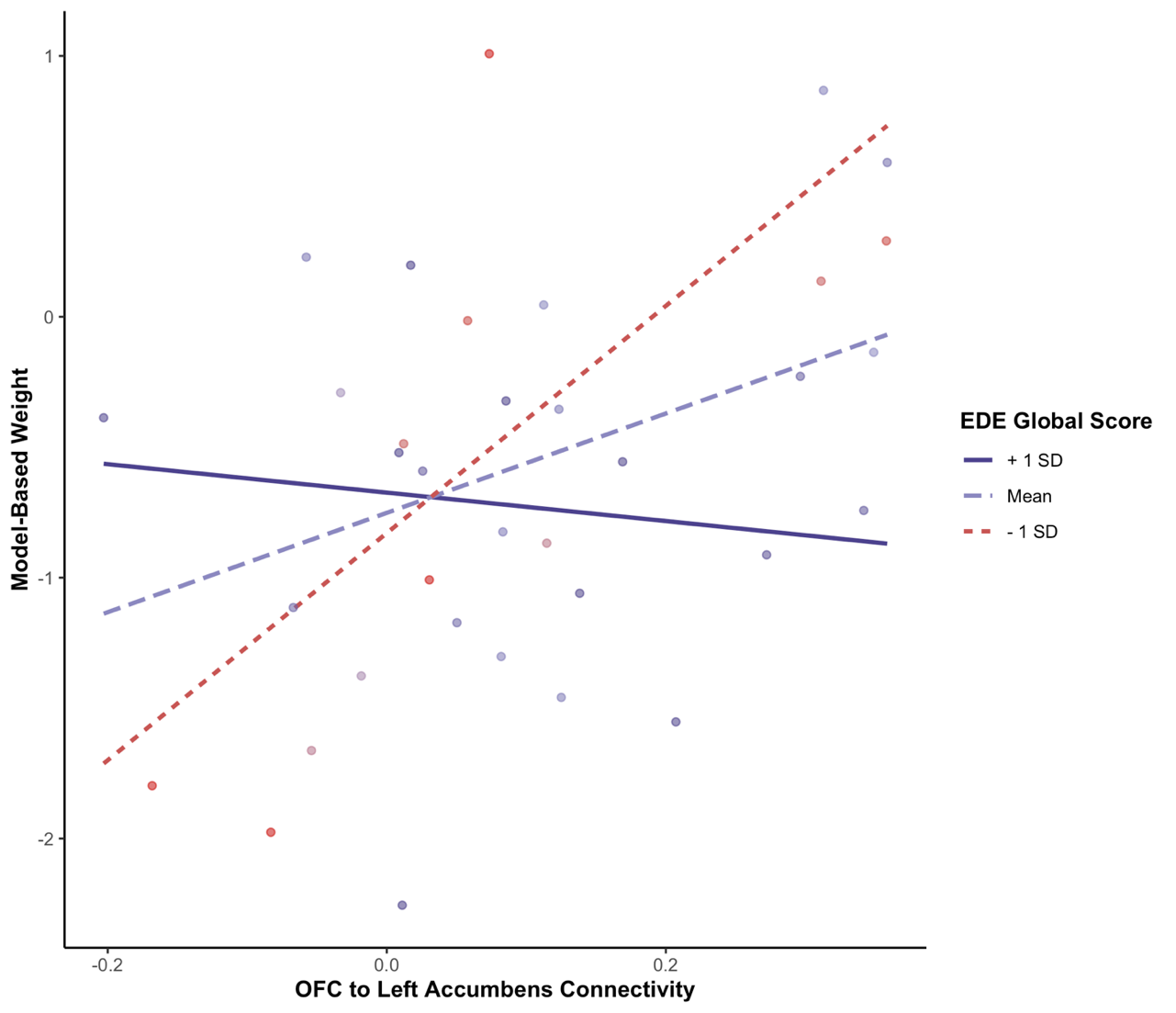

**Figure S7**. Visualization of the interaction between EDE Global Score and OFC-to-left NAcc connectivity on reward-related model-based learning in AN-R only.

| Table S7.  *Reward Model-based Weights and OFC-to-left NAcc-by-EDE Total Score Linear Regression in AN-R* | | | |  |
| --- | --- | --- | --- | --- |
| **Variable** | **Estimate** | **Standard Error** | **Statistic** | ***p*** |
| Intercept | -0.99 | 0.33 | -2.99 | **.006** |
| OFC-NAcc connectivity | 7.53 | 2.07 | 3.64 | **.001** |
| EDE Total Score | 0.06 | 0.11 | 0.56 | .579 |
| Age | -0.07 | 0.14 | -0.51 | .616 |
| BMI | -0.24 | 0.19 | -1.23 | .230 |
| WASI Full Scale IQ | 0.19 | 0.12 | 1.53 | .138 |
| OFC-NAcc connectivity x EDE Total Score | -1.92 | 0.70 | -2.74 | **.011** |

*Note*. EDE – Eating Disorder Examination; WASI – Weschler Abbreviated Scale of Intelligence.

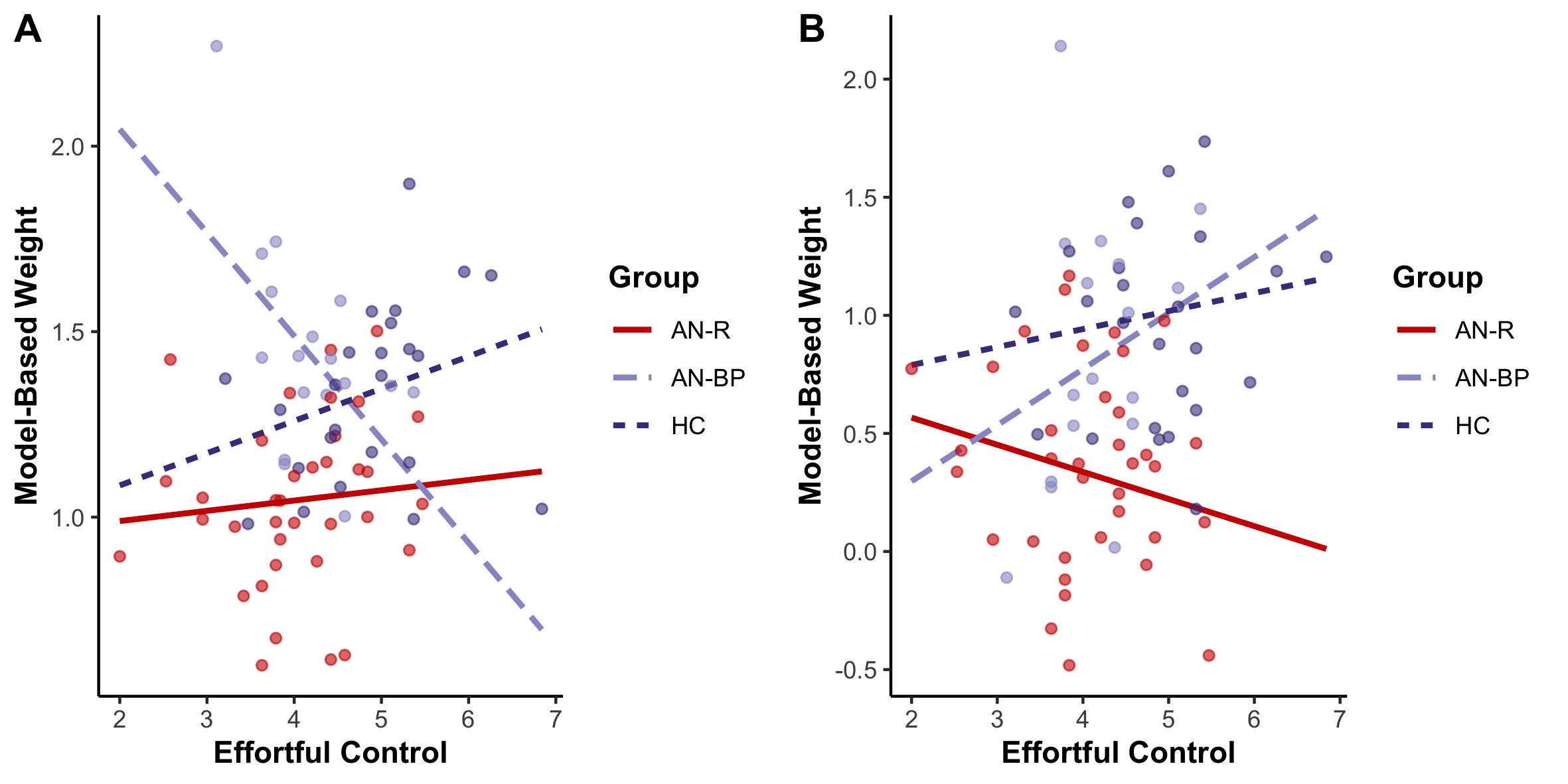

**Figure 7.** Association between model-based weights and ATQ Effortful Control scale for reward (A) and punishment (B).

| Table S8.  *Model-based Weights and Effortful Control Mixed Effects Regression* | | | |  |
| --- | --- | --- | --- | --- |
| **Variable** | **Estimate** | **Standard Error** | **Statistic** | ***p_corrected_*** |
| Intercept [AN-R] | -1.10 | 0.71 | -1.54 | 1.00 |
| AN-BP | 5.41 | 1.68 | 3.22 | .017 |
| HC | -0.25 | 1.23 | -0.21 | 1.00 |
| Condition [punishment] | 1.36 | 0.92 | 1.48 | 1.00 |
| ATQ Effortful Control | 0.11 | 0.17 | 0.62 | 1.00 |
| Age | -0.02 | 0.07 | -0.27 | 1.00 |
| BMI | 0.20 | 0.07 | 2.82 | .070 |
| WASI Full Scale IQ | -0.03 | 0.11 | -0.29 | 1.00 |
| AN-BP x Condition [punishment] | -7.94 | 2.19 | -3.63 | .006 |
| HC x Condition [punishment] | 0.03 | 1.60 | 0.02 | 1.00 |
| AN-BP x ATQ Effortful Control | -0.98 | 0.40 | -2.45 | .169 |
| HC x ATQ Effortful Control | 0.26 | 0.27 | 0.98 | 1.00 |
| Condition [punishment] x ATQ Effortful Control | -0.32 | 0.22 | -1.42 | 1.00 |
| AN-BP x Condition [punishment] x ATQ Effortful Control | 1.80 | 0.52 | 3.45 | .010 |
| HC x Condition [punishment] x ATQ Effortful Control | 0.08 | 0.35 | 0.23 | 1.00 |

*Note*. AN-R – restricting type anorexia nervosa; AN-BP – binge/purge type anorexia nervosa; HC – healthy controls; ATQ - Adult Temperament Questionnaire; WASI – Weschler Abbreviated Scale of Intelligence.

***References***

Beck, A., Steer, R., & Brown,G. (1996). *Beck Depression Inventory —Second Edition. Manual.* San Antonio, TX: The Psychological Corporation.

Carver, C. S., & White, T. L. (1994). Behavioral inhibition, behavioral activation, and affective responses to impending reward and punishment: The BIS/BAS Scales. *Journal of Personality and Social Psychology*. https://doi.org/10.1037//0022-3514.67.2.319

Cloninger, R., Svrakic, D., & Przybeck, T. (1994). The temperament and character inventory (TCI): A guide to its development and use. *Archives of General Psychiatry*.

Cooper, Z., & Fairburn, C. (1987). The eating disorder examination: A semi‐structured interview for the assessment of the specific psychopathology of eating disorders. *International Journal of Eating Disorders*, *6*(1), 1–8. https://doi.org/10.1002/1098-108X(198701)6:1<1::AID-EAT2260060102>3.0.CO;2-9

Decker, J. H., Otto, A. R., Daw, N. D., & Hartley, C. A. (2016). From Creatures of Habit to Goal-Directed Learners: Tracking the Developmental Emergence of Model-Based Reinforcement Learning. *Psychological Science*, *27*(6), 848–858. https://doi.org/10.1177/0956797616639301

Evans, D. E., & Rothbart, M. K. (2007). Developing a model for adult temperament. *Journal of Research in Personality*, *41*(4), 868–888. https://doi.org/https://doi.org/10.1016/j.jrp.2006.11.002

KAUFMAN, J., BIRMAHER, B., BRENT, D., RAO, U. M. A., FLYNN, C., MORECI, P., WILLIAMSON, D., & RYAN, N. (1997). Schedule for Affective Disorders and Schizophrenia for School-Age Children-Present and Lifetime Version (K-SADS-PL): Initial Reliability and Validity Data. *Journal of the American Academy of Child & Adolescent Psychiatry*, *36*(7), 980–988. https://doi.org/https://doi.org/10.1097/00004583-199707000-00021

Spielberger, C., Gorsuch, R., & Lushene, R. (1970). STAI Manual for the State Trait Anxiety Inventory. Palo Alto, CA:Consulting Psychologists Press.

Sutton, R. & Barto, A. (2018). Reinforcement Learning: An Introduction (2nded.). Cambridge, MA: The MIT Press.

Torrubia, R., Ávila, C., Moltó, J., & Caseras, X. (2001). The Sensitivity to Punishment and Sensitivity to Reward Questionnaire (SPSRQ) as a measure of Gray’s anxiety and impulsivity dimensions. *Personality and Individual Differences*, *31*(6), 837–862. https://doi.org/https://doi.org/10.1016/S0191-8869(00)00183-5
